# Does physical activity in childhood or adolescence predict future anxiety, depression, or wellbeing? A systematic review of 98 prospective cohort studies

**DOI:** 10.1101/2023.06.28.23292009

**Authors:** Christopher Knowles, Kyle F. Paradis, Stephen Shannon, Gavin Breslin, Angela Carlin

## Abstract

Physical activity (PA) is a modifiable health behaviour that can support and improve mental health. The tendency for activity levels to track over time suggests that through continued participation, those most active in youth may experience better long-term mental health than less active peers. Exploring the extent to which mental health benefits of child/adolescent PA persevere over time helps advocates and policy-makers determine whether PA promotion in youth constitutes effective allocation of public health resources and a viable means of reducing the global burden of common mental disorders and suboptimal wellbeing across the lifespan. This systematic review synthesised evidence for childhood/adolescent PA as a predictor of future anxiety, depression and wellbeing indicators (PROSPERO: CRD42021242555). Systematic searches of CINAHL, Medline, PsycInfo, and Scopus (conducted 27^th^ October 2022) returned 12,703 articles with 98 retained. Included: prospective, quantitative, longitudinal designs; PA measured ages 5-17 years; anxiety, depression, and/or wellbeing indicators measured at least 12 months later; generally healthy populations. Excluded: severe mental illnesses. Risk of bias and quality of evidence were assessed following the GRADE framework. Effect sizes from moderate- and high-quality studies are discussed narratively. Studies are grouped by mental health outcome, PA type, domain, and whether participants adhered to current World Health Organisation PA guidelines. PA was negatively associated with depression in 41/67 studies (61.2%); anxiety in 20/35 studies (57.1%); and positively associated with wellbeing in 25/39 studies (64.1%). Effect sizes for all outcomes were small and evidence quality low across all timeframes. Team sports were the most consistently predictive type of PA. Whether domain-specific activities or guideline (non)adherence have differential effects remains unclear. Heterogeneity was attributed to the vastness of associations tested and psychometric measures used. Current literature offers low-quality partial evidence that childhood/adolescent PA has small beneficial effects for prospective anxiety, depression and wellbeing at least 12 months later.

## Introduction

### Physical activity & mental health

There is global consensus that Physical Activity (PA) can be a viable low-risk, low-cost mental health intervention throughout the lifespan and across the world. Researchers and leading public health authorities are agreed that PA can be effective at managing depression [1], anxiety [2] and wellbeing [3]. The European Psychiatric Committee declared that PA can be at least as effective as antidepressant drugs in reducing depressive symptoms [4]. A scientific report submitted to the US Secretary of Health and Human Services found strong evidence that moderate-to-vigorous PA (MVPA) can reduce state and trait anxiety in multiple age groups [5]. The Australian Department of Health and Aged Care recommends that following their 24-hour movement guidelines, should result in better wellbeing for all age groups, with low risk of serious injury [6],[7].

### A two-continua approach to mental health

Keyes’ two-continua model recognises mental illness and wellbeing as two independent, albeit co-related, concepts existing within overall mental health [8]. Keyes’ model suggests that wellbeing is not synonymous to the absence of mental illness. Rather, individuals can and should actively pursue social, emotional, and psychological wellbeing outcomes alongside any steps taken to reduce their risk of illness [9]. An individual with high levels of wellbeing and low levels of mental illness may be described as flourishing [10],[11].

Flourishing is the epitome of mental health. A nationally representative study from the Netherlands reported that only 37% of the population were flourishing [12] inferring many people are failing to reach their potential in areas such as psychosocial functioning, occupational productivity, and healthcare utilisation [9]. Whilst up-to-date research is required to substantiate these statistics in the present day, it remains likely that sub-optimal mental health is having a greater impact than is commonly understood. Health behaviours such as PA that have potential to increase the prevalence of flourishing through management of mental illness symptomology and wellbeing support may be best placed to help build a more prosperous and healthy society.

### Anxiety & depression

The most Common Mental Disorders (CMDs) – Anxiety and Depression – are leading contributors to the global burden of disease and are becoming increasingly prevalent over time [13]. Depression and anxiety affect 280 million (23 million children/adolescents) and 301 million (58 million children/adolescents) globally [13] with one in three individuals likely to experience a CMD at some point in their lives [14]. From an economic-productivity perspective, scaling up treatments for CMDs is projected to have a combined benefit of $310 billion globally by 2030 [15]. From a health perspective, this equates to 43 million extra healthy life years [15],[16]. The investment needed to sufficiently scale-up treatments is considerable, making low-cost methods of managing CMDs including through effective PA promotion, highly desirable.

### Wellbeing

Although an historically elusive construct to define, wellbeing can be considered a state of positive feeling and meeting one’s full potential in the world [17]. Whilst CMDs consist of symptoms of mal- or sub-optimal-functioning, wellbeing consists of symptoms of positive functioning and/or positive characteristics [18]. Compared to CMDs, indicators of wellbeing have been less heavily researched thus, the economic impact of poor wellbeing is not widely known. That being said, elevated levels of wellbeing have been associated with greater productivity, more meaningful relationships and pro-social behaviour [18],[19] all of which have economic and societal implications.

### Children & adolescents

Despite evidence for the benefits of PA, global rates of inactivity remain high. In 2019, the WHO updated their Global Action Plan for PA in which they set out to achieve a 15% reduction in inactivity by 2030 [20]. On their current trajectory, it appears the WHO may fall short of that target [21],[22]. Given that resources dedicated to PA promotion on a national level are often limited, public health would benefit from a better understanding of where investment in PA promotion is likely to see the best return with regards to long-term mental health benefits. To this end, it is possible that children and adolescents are a key target population.

Whilst associations between PA and mental health in younger populations are less consistently observed than in adults, previous reviews have concluded there is partial evidence for a causal link between child/adolescent PA, lower levels of mental illness, and greater wellbeing [23]-[25]. One important causal pathway through which child/adolescent PA may support long-term mental health is in the tendency for PA levels to track across the lifespan [26],[27]. Whilst rates of activity generally decrease as we age, those most active early in life tend to remain the most active as adults [27]. This means interventions successful at upscaling PA in children and adolescents not only have potential to benefit young people in the short-term but may improve future rates of participation and subsequently, future mental health [28]. Such an outcome would constitute an effective use of public health resources, particularly for young people – a high risk group due to high rates of inactivity and the early age of onset of symptoms of mental illness [29]-[31].

A comprehensive systematic review of the literature investigating associations between child/adolescent PA and future mental health outcomes has not been conducted. To better understand these longitudinal associations, a review should consider relationships measured over lengthy timeframes with outcomes pertaining to both mental illness and wellbeing. A time interval of at least one year between first PA measurement and prospective mental health outcomes represents a significant window in which to capture psychological developmental changes and would strengthen understanding of how long associations persist over time [32],[33].

### The current study

The aim of the present systematic review was to synthesise evidence for a longitudinal relationship between PA participation in youth and prospective mental health outcomes measured at least one year later. To align with Keyes’ two-continua model, the review included measures of mental illness (i.e., anxiety, depression) and indicators of social, emotional and psychological wellbeing. The review has been reported in line with the recently updated Preferred Reporting Items for Systematic Reviews and Meta-Analyses (PRISMA) guidelines [34]. The following review questions were written *a priori*:

1. Can PA performed in youth reduce depressive disorders/symptoms at least 12 months later?
2. Can PA performed in youth reduce anxiety disorders/symptoms at least 12 months later?
3. Can PA performed in youth improve indicators of wellbeing at least 12 months later?

To offer the most holistic insight of associations observed to date we reviewed PA in its broadest sense, “any form of bodily movement performed by skeletal muscles that results in an increase in energy expenditure” [20],[35]. To make findings useful in a real-world context, we then sorted the PA performed into subgroups were possible. Public health messaging consistently emphasises the importance of children/adolescents achieving WHO guidelines of 60minutes MVPA per day. Additionally, effects are often dependent on the type of activity (i.e., individual or group-based) [36],[37] and the life domain it is performed in (i.e., leisure, occupational/school-based, transport-based, household) [38],[39]. Therefore, as secondary lines of enquiry, we also aimed to synthesise evidence for the effects of (non)adherence to current WHO MVPA guidelines, individual versus group-based activities, and PA in different life domains where possible.

## Materials & methods

The review protocol was published on PROSPERO on 15^th^ March 2021 prior to any formal database searches and can be accessed via the registration number: CRD42021242555.

### Eligibility criteria

#### Participants

Included studies were restricted to those published in English using generally healthy populations [38]. Studies were also restricted to those that collected PA data from school-aged children/adolescents between the ages of 5 and 17 years. Many PA interventions in youths are carried out in-school so using the school years as our upper and lower age thresholds seemed a sensible way to define the target population. Further, the WHO propose PA guidelines for those aged 5 to 17 years old so using these age thresholds best facilitated a review of the effects of adherence to the current youth guidelines. If studies included samples where some participants were within this range and some not (e.g., 15 to 20 years old), they were included if the mean age of the sample fell within the target range. At no point did review authors have access to information that could be used to personally identify participants.

#### Study length

When first conceptualising the review, the original aim was to capture studies that followed participants as they moved from one key life stage to another. In principle, this was to help answer whether more active young people make more mentally healthy adults. Operationally, this proved difficult and came with the risk that a three-year-long study may be excluded if it were performed between the ages of 6 and 9 (i.e., all within childhood) yet a one-year-long study would be included if it followed participants from age 17 to 18 thus covering what arguably denotes a transition from adolescence into adulthood. Ongoing debate regarding the age at which one life stage ends and another begins added a further layer of complexity to achieving this research aim and made robust methodology elusive [40]. The most inclusive and methodologically coherent approach to achieve our research aim was to target studies that analysed mental health data collected at least 12 months after initial PA measurement. Whilst this meant that there would be studies included that did not follow participants as they moved through different life stages, it would offer a more complete insight into the perseverance of PA-related mental health benefits over time and limit the chance that important studies were overlooked.

#### Measures

Short-term recall questionnaires typically gather information pertaining to the past few weeks or months whilst quantitative history questionnaires probe memories from several years ago or across an entire lifespan [41]. To minimise the risk of recall bias, measures of PA had to constitute short-term recall (i.e., PA within the last year) or be collected by wearable devices.

Included studies investigated anxiety, depression, and/or indicators of wellbeing. As the two-continua model views mental illness and wellbeing as independent concepts, measures were excluded if they combined items targeting depression/anxiety and wellbeing into one overall mental health score. The focus was on CMDs rather than severe mental illnesses (e.g., suicidal ideation, psychosis) due to their comparatively greater contribution to the overall global burden of disease [13].

An operational definition of flourishing identifies 10 symptoms said to represent the mirror opposite of CMDs [18]. Competence, engagement, meaning, and positive relationships represent elements of positive functioning whilst emotional stability, optimism, positive emotions, resilience, self-esteem, and vitality represent positive characteristics. Including all 10 factors in a single review would be unwieldy. Instead, three items were identified through group discussion that either represent or heavily contribute to social, emotional, and psychological wellbeing as per the two-continua model and the definition of flourishing discussed. Positive emotions (e.g., happiness, life satisfaction) are essential criteria for flourishing hence, these measures qualified as indicators of emotional wellbeing [18]. Positive relationships (e.g., social competence, peer-relations) and self-esteem represented indicators of (1) positive functioning/social wellbeing, and (2) positive characteristics/psychological wellbeing, respectively. Self-esteem was selected as a requisite for flourishing over other positive characteristics given its importance to schools and other key stakeholders.

### Search strategy, study selection and data collection

Systematic searches of CINAHL Complete, Medline, PsycInfo, and Scopus were conducted from inception to 27^th^ October 2022. Key words used to capture studies of CMDs were derived from DSM-5 definitions of anxiety/depressive disorders. Huppert and So’s operational definition of flourishing [18] and Keyes’ two-continua model [8] were used to identify key words relating to positive emotions (emotional wellbeing), positive relationships (social wellbeing) and self-esteem (psychological wellbeing). Reference lists of similar existing reviews were manually searched as were authors own libraries. Search results were uploaded to online systematic review software Covidence where eligibility criteria were visibly displayed above each article. Authors confirmed each point sequentially to determine eligibility. Full search strategies for all databases and a succinct version of the eligibility criteria can be found in the published review protocol.

Articles first underwent title and abstract screening by a single reviewer removing those that were clearly irrelevant to the focus of the review. It is not essential that this stage be carried out by two reviewers [42] nevertheless, 1270 excluded studies (10%) were screened again by a second reviewer. Duel screening a random subset of excluded studies allowed reviewers to estimate the number of studies mistakenly omitted and assess inter-rater reliability. Next, two authors independently reviewed the full texts of remaining articles to determine which should be included in final analysis. Disputes were resolved through discussion with a third and/or fourth reviewer. Data extraction was conducted by one reviewer using a bespoke data extraction template.

### Data items

Data were extracted on study characteristics including authors, location, study design, the name of the dataset used (if applicable), and the data collection schedule, as well as the age, number, and sex/gender split of participants. Further information included: the source of PA and mental health data (e.g., self-report, accelerometers); measures used; association(s) tested; confounding variables; analysis performed; and results. Information provided by study authors and/or the wording of specific items were used to categorise the type and domain of PA, and whether participants adhered to PA guidelines. Further information was extracted to aid quality of evidence and risk of bias assessment, explained in detail below.

### Evidence quality & risk of bias

A quality of evidence and risk of bias assessment was made for each study by two independent reviewers using the framework proposed by the GRADE Working Group [43]. Evidence was graded as high, moderate, low, or very low. The items used to investigate certainty of evidence, sources of heterogeneity, and risk of bias have been published elsewhere [43].

### Synthesis methods

Included studies were heterogeneous with regards to the timespans covered and associations tested. After discussion, authors agreed the review was best suited to an exploratory analysis of potential sources of heterogeneity through narrative synthesis to offer more nuanced conclusions. Results were therefore synthesized according to the Synthesis Without Meta-Analysis (SWiM) reporting guidelines [44].

Studies reporting statistically significant associations at the 0.05 level were considered as providing evidence of the direction of an effect. The total number of studies reporting evidence (or lack thereof) an effect were summed and grouped by mental health outcome and length of follow-up in years to provide an overview of the literature as a whole and the timescales across which childhood/adolescent PA was statistically significantly predictive of future mental health. This approach has been taken in existing reviews in other areas of public health[45]. However, the large sample sizes often used meant it was not surprising to find a considerable number of statistically significant associations. To avoid over-reliance on statistical significance as evidence of an effect and the risk that inappropriate conclusions would be drawn from low quality evidence; effect sizes reported in moderate- and high-quality studies were interpreted and discussed using thresholds proposed in existing research [46],[47].

Finally, the main analysis was supplemented with important contextual information on studies investigating type/domain of activity and guideline adherence to strengthen depth of understanding. Summary of Findings (SoF) tables for each secondary review question include a brief explanation of the associations tested and effect sizes reported in each study (see supporting information). Again, this approach has been used in existing reviews [48].

All studies were eligible for data synthesis provided they did not replicate research from other studies using the same dataset. This was to limit the risk that data from a single source skewed conclusions drawn [49]. Supporting information (File S2) has been provided on the study modifiers used to differentiate between studies using the same datasets to ensure each made a unique contribution to knowledge. Modifiers related to Participants, Exposure, Outcomes and Study Design.

## Results

### Study selection

After removal of duplicates, systematic searches returned 12,703 articles to be screened by title and abstract. A total of 496 articles underwent full text review. Conflicts were resolved and data was extracted from 100 studies. A further two studies were removed^(74, 86)^ post-extraction as they replicated research conducted by other included studies using the same data-source. Supporting information has been provided detailing the decision-making process that led to the exclusion of these studies (Supporting Table S1). This left 98 studies eligible for synthesis.

Of the 1270 excluded studies reviewed by a second reviewer, 25 (2.3%) were retained for full text review of which only 1 (0.1%) was included in final analysis. This speaks to the rigor with which title and abstract screening was carried out indicating the use of a single reviewer at this stage did not meaningfully detract from review reliability. A detailed report of the number of articles returned per database and the reasons for exclusion are presented in the PRISMA flowchart (Fig 1).

**Figure 1:**
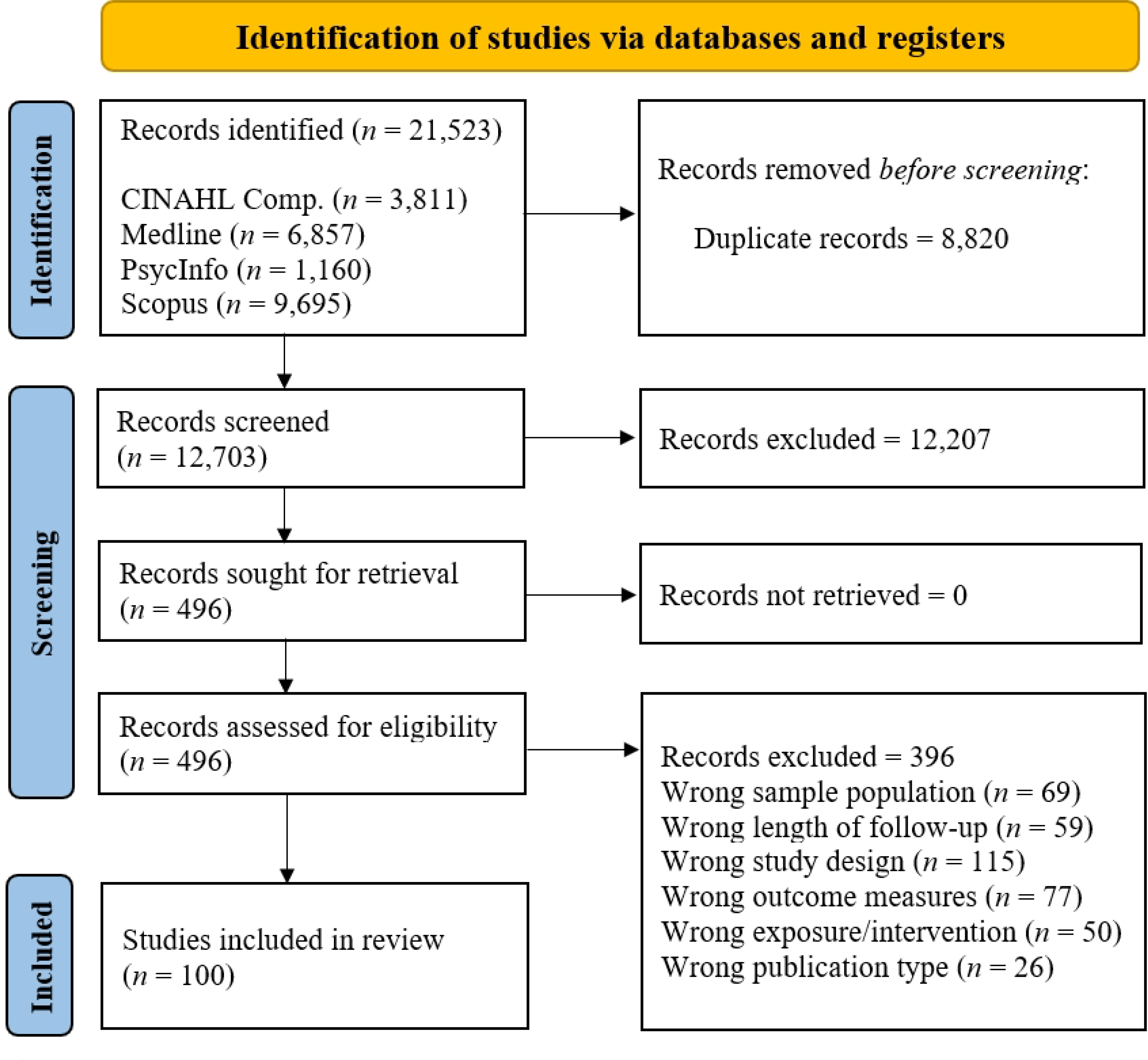
PRISMA flowchart

### Study characteristics

*Table 1* provides an index of all original 100 articles. Of the 98 articles included in the synthesis, there were 94 observational cohort studies (95.9%) and four randomized trials^(8, 49, 53, 54)^. Studies were conducted throughout North America (*n* = 48), Europe (*n* = 37), Australia (*n* = 11) and Asia (*n* = 2). Secondary analysis was conducted by 81 studies using data from 53 different longitudinal cohort study projects. The most heavily used data source was the National Longitudinal Study of Adolescent to Adult Health (Add Health) used in 10 separate studies^(10, 15, 19, 21, 25, 44, 48, 61, 96, 98)^ followed by the Nicotine Dependence in Teens Study (NDIT)^(5, 14, 39, 50, 63)^.

**Table 1:** Index of all included studies and the mental health outcomes they investigated

| ID | Authors (Date of Publication) | Mental Illness |  | Wellbeing |  |  | ID | Authors (Date of Publication) | Mental Illness |  | Wellbeing |  |  |
| --- | --- | --- | --- | --- | --- | --- | --- | --- | --- | --- | --- | --- | --- |
|  |  | Dep. | Anx. | Psych. | Soc. | Emo. |  |  | Dep. | Anx. | Psych. | Soc. | Emo. |
| 1 | Adachi & Willoughby (2014) |  |  | ✓ |  |  | 30 | Gunnell et al. (2016a) | ✓ | ✓ |  | ✓ |  |
| 2 | Agans et al. (2017) | ✓ |  |  |  |  | 31 | Gunnell et al. (2016b) | ✓ | ✓ |  |  |  |
| 3 | Ahn et al. (2018) |  | ✓ |  | ✓ |  | 32 | Hamer et al. (2020) | ✓ |  |  |  |  |
| 4 | Ames & Leadbeater (2018) | ✓ |  |  |  |  | 33 | Herman et al. (2010) |  |  |  | ✓ |  |
| 5 | Ashdown-Franks et al. (2017) |  | ✓ |  |  |  | 34 | Hoegh-Poulsen et al. (2016) | ✓ |  |  |  |  |
| 6 | Barber et al. (2001) | ✓ | ✓ | ✓ |  |  | 35 | Howie et al. (2016) | ✓ | ✓ |  |  |  |
| 7 | Bedard et al. (2020) |  |  |  | ✓ |  | 36 | Howie et al. (2020) | ✓ | ✓ |  |  |  |
| 8 | Bell et al. (2019) |  | ✓ | ✓ | ✓ | ✓ | 37 | Isaksson et al. (2020) | ✓ | ✓ |  |  |  |
| 9 | Birkeland et al. (2009) | ✓ |  |  |  |  | 38 | Jerstad et al. (2010) | ✓ |  |  |  |  |
| 10 | Bohr et al. (2019) | ✓ |  |  |  |  | 39 | Jewett et al. (2014) | ✓ |  |  |  |  |
| 11 | Brière et al. (2020) | ✓ | ✓ |  |  |  | 40 | Kalimo & Vuori (1990) |  |  |  |  | ✓ |
| 12 | Brière et al. (2018) | ✓ | ✓ |  |  |  | 41 | Kandola et al. (2020a) | ✓ |  |  |  |  |
| 13 | Brunes et al. (2015) | ✓ | ✓ |  |  | ✓ | 42 | Kandola et al. (2020b) |  | ✓ |  |  |  |
| 14 | Brunet et al. (2013) | ✓ |  |  |  |  | 43 | Kaseva et al. (2016) | ✓ |  |  |  |  |
| 15 | Carter et al. (2015) | ✓ |  |  |  |  | 44 | Kort-Butler & Hagewen (2011) |  |  | ✓ |  |  |
| 16 | Carvajal (2012) | ✓ |  |  |  |  | 45 | Mahoney & Vest (2012) | ✓ | ✓ | ✓ | ✓ | ✓ |
| 17 | Clark et al. (2007) | ✓ |  |  |  |  | 46 | Marsh & Kleitman (2003) |  |  | ✓ |  |  |
| 18 | Colman et al. (2014) | ✓ |  |  |  |  | 47 | McKercher et al. (2014) | ✓ |  |  |  |  |
| 19 | Daniels & Leaper (2006) |  |  | ✓ |  |  | 48 | McPhie & Rawana (2015) | ✓ |  |  |  |  |
| 20 | Denalt et al. (2009) | ✓ |  |  |  |  | 49 | Motl (2004) | ✓ |  |  |  |  |
| 21 | Deshpande et al. (2020) | ✓ | ✓ |  |  |  | 50 | Murray et al. (2021) | ✓ | ✓ |  |  |  |
| 22 | Doré et al. (2020) |  |  | ✓ | ✓ | ✓ | 51 | Nigg et al. (2021) |  | ✓ |  | ✓ |  |
| 23 | Doré et al. (2019) |  |  | ✓ | ✓ | ✓ | 52 | Noordstar et al. (2016) |  |  | ✓ |  |  |
| 24 | Duncan et al. (2021) | ✓ | ✓ | ✓ | ✓ | ✓ | 53 | Olive et al. (2019) | ✓ |  |  |  |  |
| 25 | Easterlin et al. (2019) | ✓ | ✓ |  |  |  | 54 | Omorou et al. (2016) |  |  |  | ✓ |  |
| 26 | Fløtnes et al. (2011) | ✓ | ✓ |  |  |  | 55 | Patte et al. (2020) | ✓ |  |  |  |  |
| 27 | Gómez-Baya et al. (2018) |  |  |  |  | ✓ | 56 | Perron et al. (2012) | ✓ |  |  |  |  |
| 28 | Gómez-Baya et al. (2020) | ✓ |  |  |  |  | 57 | Piché et al. (2019) | ✓ |  |  |  |  |
| 29 | Graupensperger et al. (2021) | ✓ | ✓ |  | ✓ |  | 58 | Rangul et al. (2012) | ✓ |  |  |  | ✓ |
|  |  | Dep. | Anx. | Psych. | Soc. | Emo. |  |  | Dep. | Anx. | Psych. | Soc. | Emo. |
| 59 | Raudsepp & Neissaar (2012) | ✓ |  |  |  |  | 80 | Toseeb et al. (2014) | ✓ |  |  |  |  |
| 60 | Raudsepp & Vink (2019) | ✓ |  |  |  |  | 81 | Van Dijk et al. (2016) | ✓ |  | ✓ |  |  |
| 61 | Rees & Sabia (2010) | ✓ |  | ✓ |  |  | 82 | Visser et al. (2020) |  |  | ✓ | ✓ |  |
| 62 | Rongen et al. (2020) |  |  |  | ✓ |  | 83 | Wagnsson et al. (2014) |  |  | ✓ |  |  |
| 63 | Sabiston et al. (2016) | ✓ |  |  |  |  | 84 | Wang et al. (2017) | ✓ |  |  |  |  |
| 64 | Sagatun et al. (2007) |  | ✓ |  | ✓ |  | 85 | Wiles et al. (2008) |  | ✓ |  |  |  |
| 65 | Sánchez-Oliva et al. (2020) | ✓ |  |  |  | ✓ | 86 | Wu et al. (2017) | ✓ | ✓ |  |  |  |
| 66 | Schmalz et al. (2007) |  |  | ✓ |  |  | 87 | Wu et al. (2018) | ✓ |  |  |  |  |
| 67 | Schumacher-Dimech & Seiler (2011) |  | ✓ |  |  |  | 88 | You et al. (2021) | ✓ |  |  |  | ✓ |
| 68 | Seeley et al. (2009) | ✓ |  |  |  |  | 89 | Wilson et al., (2022) |  |  |  |  | ✓ |
| 69 | Shin & You (2013) |  |  |  |  | ✓ | 90 | Redig et al., (2022) | ✓ |  |  |  |  |
| 70 | Slutzky & Simpkins (2009) |  |  | ✓ |  |  | 91 | Kandola et al., (2022) |  | ✓ |  |  |  |
| 71 | Slykerman et al. (2020) | ✓ |  |  |  |  | 92 | Barth Vedøy et al., (2021) |  |  | ✓ | ✓ | ✓ |
| 72 | Spreitzer (1994) |  |  | ✓ |  |  | 93 | O'Flaherty et al., (2022) | ✓ | ✓ |  | ✓ |  |
| 73 | Stavrakakis et al. (2012) | ✓ |  |  |  |  | 94 | Jetten et al., (2022) | ✓ | ✓ |  |  |  |
| 74 | Stavrakakis et al. (2013a) | ✓ |  |  |  |  | 95 | Harbec et al., (2022) | ✓ | ✓ |  |  |  |
| 75 | Stavrakakis et al. (2013b) | ✓ |  |  |  |  | 96 | García-Hermoso et al., (2022) | ✓ |  |  |  |  |
| 76 | Stein et al. (2007) |  |  | ✓ | ✓ |  | 97 | Buchan et al., (2021) | ✓ | ✓ |  |  |  |
| 77 | Ströhle et al. (2007) | ✓ | ✓ |  |  |  | 98 | Iverson et al., (2021) | ✓ |  |  |  |  |
| 78 | Suetani et al. (2017) | ✓ | ✓ |  |  |  | 99 | White-Gosselin et al, (2022) | ✓ | ✓ |  |  |  |
| 79 | Sund et al. (2011) | ✓ |  |  |  |  | 100 | Latina et al., (2022) | ✓ |  | ✓ |  |  |

The remaining 17 studies appeared to use primary data collected by the lead author or their research partners^(16, 20, 27, 28, 38, 59, 60, 62, 65, 66, 67, 68, 82, 83, 92, 94, 100)^. The most heavily researched mental health outcome was depression, investigated in 67/98 studies (68.4%) compared to 35/98 (35.7%) investigating anxiety and 39/98 (39.8%) investigating wellbeing. A quality of evidence assessment was conducted for each study: High (*n* = 4), Moderate (*n* = 10), Low (*n* = 59), or Very Low (*n* = 25). Overall, the quality of evidence investigating the association between childhood/adolescent PA and mental health outcomes at least 12 months later was low for anxiety, low for depression, and low for indicators of wellbeing.

Studies were published from 1990–2022. There were noticeable upticks in publications in 2007, 2016, and 2020 signalling a gradual but consistent increase in research output in the area (Fig 2). A total of 75/98 (76.5%) articles were published from 2012-2022 and around half (48/98, 49.0%) were from 2017 onward meaning most of the research reviewed was recently published and up to date at the time of writing. Evidence spanned 65 years with studies availing of PA and mental health data collected from 1955^(40)^ to 2020^(90)^. Moderating effects of time are discussed herein. All studies used data collected before the outbreak of the Covid-19 pandemic. It remains to be seen whether young people engaging in PA before Covid-19 experienced mental health benefits upon emergence from the pandemic compared to their previously less active peers.

**Figure 2:**
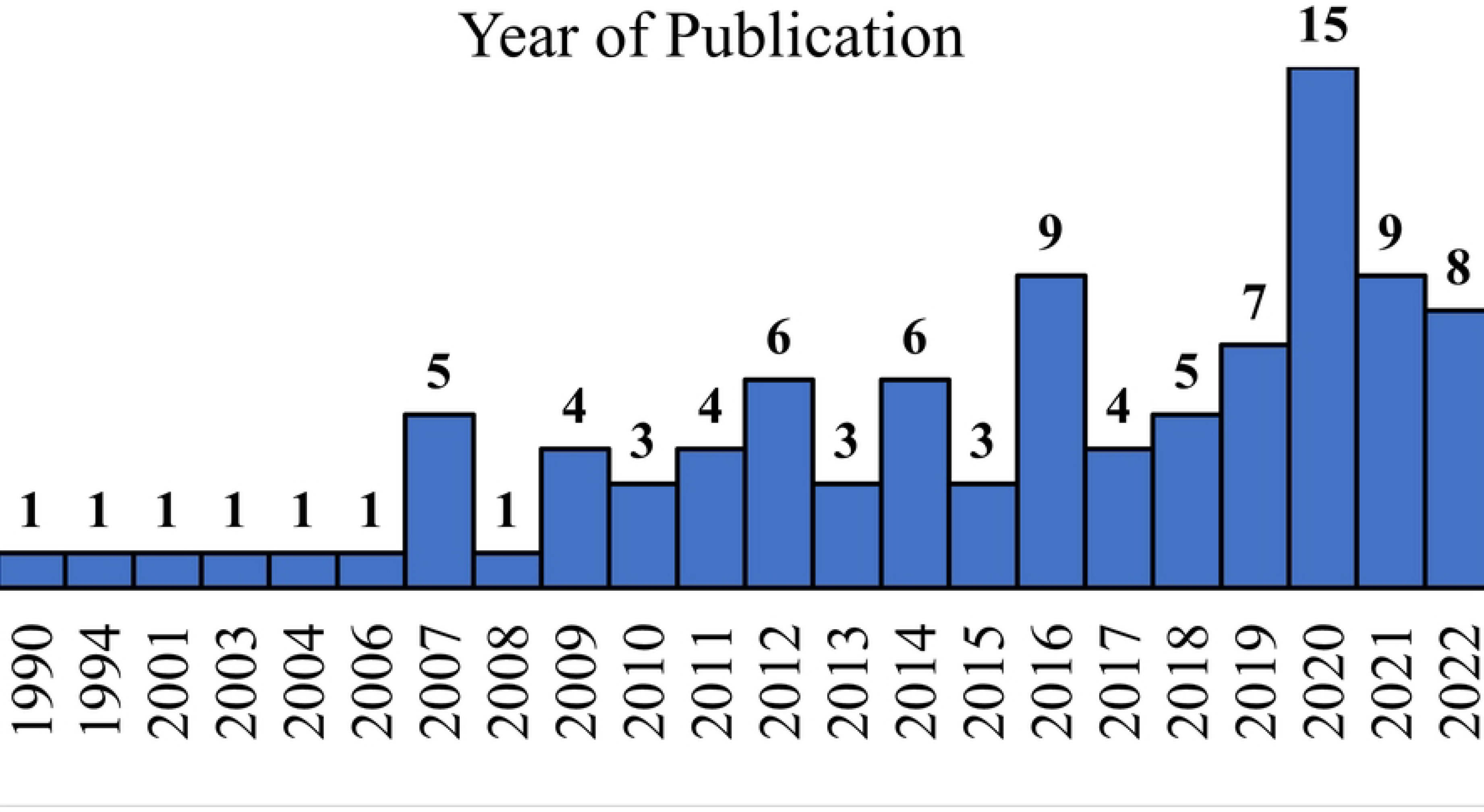
Year of publication

Self-report measures of PA were used in 88 studies, 11 used mechanical device-measured data^(3, 8, 32, 41, 42, 65, 71, 80, 81, 82, 92)^. One study used details of a football academy’s weekly training schedule as its PA data source^(62)^ and one study used trained observers reports of children in a PE class^(53)^. All mental health outcomes were self-reported other than in one study using national medical databases to determine rates of clinical diagnoses of internalizing disorders^(87)^. A key for all acronyms used for mental health measures can be found at the bottom of Tables 2, 3, and 4, and in the supporting information. Studies used 23 different validated measures of depression of which the CES-D [50] was the most popular with 20/67 depression studies (30.0%) using a version of the scale^(2, 10, 12, 15, 16, 21, 24, 25, 34, 48, 49, 55, 59, 60, 61, 71, 81, 96, 97, 100)^. There were also 19 validated measures of anxiety and 15 validated scales measuring indicators of wellbeing. The most common measure of anxiety symptoms was the SDQ Emotional Symptoms subscale [51] used in 6/35 (17.1%) of anxiety studies^(3, 8, 51, 64, 85, 91)^.

**Table 2:** Studies investigating associations between physical activity and depression

| ID | Dataset | Participants<br>(n) | Age<br>(Baseline) | Age<br>(Follow-Up) | Study Length<br>(Years) | Measure (Subscale) | Evidence of<br>Association | Quality of<br>Evidence |
| --- | --- | --- | --- | --- | --- | --- | --- | --- |
| 90 | NCDS | 18,555 | 7 | 62 | 55 | *Malaise (depression) | Y | High |
| 87 | CLASS | 4861 | 10 | 18 | 8 | MSID; CIHIDAD | Y | High |
| 61 | Add Health | 13,426 | 12-17 | 13-18 | 1 | *CES-D | Y | Moderate |
| 96 | Add Health | 7069 | 12-17 | 33-39 | 21 | *CES-D; Diagnosis | Y | Moderate |
| 41 | ALSPAC | 4257 | 12 | 18 | 6 | *CIS | Y | Moderate |
| 63 | NDIT | 860 | 12 | 20 | 8 | *MDI | Y | Moderate |
| 39 | NDIT | 853 | 12 | 20 | 8 | *MDI | Y | Moderate |
| 53 | LOOK | 821 | 8 | 12 | 4 | *CDI | N | Moderate |
| 99 | FLUPPY | 785 | 6 | 10 | 4 | *PBQ; *CBC | Y | Moderate |
| 50 | NDIT | 733 | 12 | 20 | 8 | *MDI | N | Moderate |
| 97 | COMPASS | 23,395 | 14-18 | 15-19 | 1 | *CES-D | Y | Low |
| 10 | Add Health | 10951 | 12-17 | 29 | 13 | *CES-D | Y | Low |
| 25 | Add Health | 4888 | 12-17 | 24-32 | 13 | Diagnosis; *CES-D | Y | Low |
| 49 | TEENS | 4594 | 12 | 14 | 2 | *CES-D | Y | Low |
| 29 | LSAC | 3956 | 12 | 16 | 4 | *SMFQ | Y | Low |
| 93 | LSAC | 3885 | 13 | 15 | 2 | *SMFQ | N | Low |
| 48 | Add Health | 3676 | 15 | 28 | 13 | *CES-D | Y | Low |
| 43 | CRYF | 3596 | 9-18 | 35-50 | 32 | *MDI | Y | Low |
| 78 | MUSP | 3493 | 14 | 21 | 7 | *CIDI | Y | Low |
| 24 | COMPASS | 3173 | 14-18 | 15-19 | 1 | *CES-D | N | Low |
| 77 | EDSP | 2548 | 14-24 | 22-32 | 8 | *CIDI | Y | Low |
| 32 | MCS | 2416 | 7 | 14 | 7 | *SMFQ | Y | Low |
| 55 | COMPASS | 2292 | 15-17 | 16-18 | 1 | *CES-D | N | Low |
| 73 | TRAILS | 2230 | 11 | 16 | 5 | *YSR; *CBC | Y | Low |
| 21 | Add Health | 2197 | 12-17 | 24-32 | 14 | *CES-D | N | Low |
| 79 | YAMHS | 2175 | 14 | 15 | 1 | *SMFQ | Y | Low |
| 88 | KCYPS | 2092 | 13 | 15 | 2 | *KEBPS | Y | Low |
| 26 | HUNT | 2000 | 12-17 | 17-19 | 5 | *HSCL | N | Low |
| 58 | HUNT | 1869 | 13-19 | 23-31 | 10 | *CONOR-MHI | Y | Low |
| 35 | Raine | 1679 | 5 | 20 | 15 | *DASS-21 | Y | Low |
| 47 | CDAH | 1630 | 12-15 | 29-35 | 20 | *CIDI | N | Low |
| 34 | VestLiv | 1589 | 14-15 | 20-21 | 6 | *CES-D | Y | Low |
| 57 | QLSCD | 1516 | 7 | 8 | 1 | *SBQ | Y | Low |
| 17 | RELACHS | 1513 | 11-14 | 13-16 | 2 | *SMFQ | N | Low |
| 11 | QLSCD | 1492 | 6 | 12 | 6 | *SBQ; *PBQ | Y | Low |
| 95 | QLSCD | 1438 | 5 | 10 | 5 | *SBQ | Y | Low |
| 37 | SALVe | 1428 | 14 | 17 | 3 | *DSRS | Y | Low |
| 13 | HUNT | 1417 | 12-13 | 17-19 | 5 | *HSCL | N | Low |
| 75 | TRAILS | 1396 | 14 | 19 | 5 | *CIDI | N | Low |
| 56 | QLSCD | 1250 | 8 | 10 | 2 | *PBQ; *CBC | Y | Low |
| 31 | REAL | 1160 | 12-15 | 17-20 | 11 | *CDI | N | Low |
| 65 | UP&DOWN | 1099 | 8-18 | 10-20 | 2 | *PANAS | N | Low |
| 84 | CAB | 1065 | 12-15 | 14-17 | 2 | *HSCL | Y | Low |
| 100 | SSP | 998 | 12-14 | 13-15 | 1 | *CES-D | N | Low |
| 9 | NLHB | 924 | 13 | 23 | 10 | 7-items: depressed mood | N | Low |
| 71 | ABC | 871 | 11 | 16 | 5 | *CES-D | N | Low |
| 14 | NDIT | 816 | 12 | 20 | 8 | *MDI | N | Low |
| 80 | ROOTS | 736 | 15 | 18 | 3 | *SMFQ; *SADSSAC | N | Low |
| 4 | VHYS | 662 | 12-18 | 22-29 | 10 | *BCFPI | Y | Low |
| 28 | n/a (Primary Data) | 525 | 13-14 | 16-17 | 3 | *CDI | Y | Low |
| 38 | n/a (Primary Data) | 496 | 11-15 | 17-21 | 6 | *SADSSAC | Y | Low |
| 12 | NANS | 17550 | 12-15 | 13-16 | 1 | *CES-D | N | Very Low |
| 15 | Add Health | 6504 | 12-17 | 18-26 | 7 | *CES-D | Y | Very Low |
| 2 | 4-H | 5961 | 13 | 21 | 8 | *CES-D | Y | Very Low |
| 98 | Add Health | 2353 | 12-17 | 24-32 | 14 | *Diagnosis; 1-item: depression | N | Very Low |
| 36 | Raine | 1628 | 8 | 22 | 14 | *DASS-21 | Y | Very Low |
| 18 | NPHS | 1137 | 16-17 | 18-23 | 6 | *CIDI | Y | Very Low |
| 45 | PSID | 1115 | 12-18 | 18-24 | 6 | *KPDS | N | Very Low |
| 30 | MATCH | 932 | 9-13 | 13-17 | 4 | *PedsQL | N | Very Low |
| 6 | MSLT | 900 | 15 | 24 | 9 | 3-items: depressed mood | N | Very Low |
| 16 | Missing | 744 | 12-13 | 13-14 | 1.5 | *CES-D | Y | Very Low |
| 94 | n/a (Primary Data) | 558 | 12-17 | 13-18 | 1 | *HADS | N | Very Low |
| 68 | n/a (Primary Data) | 479 | 16 | 20 | 4 | *SADSSAC | Y | Very Low |
| 20 | n/a (Primary Data) | 362 | 13 | 16 | 3 | *CDI | N | Very Low |
| 59 | n/a (Primary Data) | 277 | 12 | 15 | 3 | *CES-D | Y | Very Low |
| 60 | n/a (Primary Data) | 173 | 13 | 15 | 2 | *CES-D | Y | Very Low |
| 81 | GOALS | 158 | 14 | 15 | 1 | *CES-D | N | Very Low |

**Table 3:** Studies investigating associations between physical activity and anxiety

| ID | Dataset | Participants<br>(n) | Age<br>(Baseline) | Age<br>(Follow-Up) | Study Length<br>(Years) | Measure (Subscale) | Evidence of<br>Association | Quality of<br>Evidence |
| --- | --- | --- | --- | --- | --- | --- | --- | --- |
| 87 | CLASS | 4861 | 10 | 18 | 8 | MSID; CIHIDAD | Y | High |
| 8 | AHEAD | 673 | 13 | 15-16 | 3 | *SDQ (Emotional Symptoms) | Y | High |
| 3 | MCS | 6153 | 7 | 11 | 4 | *SDQ (Emotional Symptoms) | N | Moderate |
| 91 | MCS | 4599 | 14 | 17 | 3 | *SDQ (Emotional Symptoms) | Y | Moderate |
| 99 | FLUPPY | 785 | 6 | 10 | 4 | *TRF | Y | Moderate |
| 50 | NDIT | 733 | 12 | 20 | 8 | CCHS | Y | Moderate |
| 97 | COMPASS | 23,395 | 14-18 | 15-19 | 1 | *GAD-7 | Y | Low |
| 25 | Add Health | 4888 | 12-17 | 24-32 | 13 | Clinical Diagnosis | Y | Low |
| 42 | ALSPAC | 4257 | 12 | 18 | 6 | *CIS | Y | Low |
| 29 | LSAC | 3956 | 12 | 16 | 4 | *SCAS | Y | Low |
| 93 | LSAC | 3885 | 13 | 15 | 2 | *PedsQL (Emotional) | Y | Low |
| 78 | MUSP | 3493 | 14 | 21 | 7 | *CIDI | N | Low |
| 24 | COMPASS | 3173 | 14-18 | 15-19 | 1 | *GAD-7 | N | Low |
| 77 | EDSP | 2548 | 14-24 | 22-32 | 8 | *CIDI | Y | Low |
| 64 | HUBRO | 2433 | 15-16 | 18-19 | 3 | *SDQ (Emotional Symptoms) | Y | Low |
| 21 | Add Health | 2197 | 12-17 | 24-32 | 14 | Clinical Diagnosis | N | Low |
| 88 | KCYPS | 2092 | 13 | 15 | 2 | *KEBPS | Y | Low |
| 26 | HUNT | 2000 | 12-17 | 17-19 | 5 | *HSCL | N | Low |
| 58 | HUNT | 1869 | 13-19 | 23-31 | 10 | *CONOR-MHI | Y | Low |
| 35 | Raine | 1679 | 5 | 20 | 15 | *DASS-21 | N | Low |
| 11 | QLSCD | 1492 | 6 | 12 | 6 | *SBQ; *PBQ | Y | Low |
| 85 | n/a (Secondary Data) | 1446 | 11-14 | 12-15 | 1 | *SDQ (Emotional Symptoms) | Y | Low |
| 95 | QLSCD | 1438 | 5 | 10 | 5 | *SBQ | Y | Low |
| 37 | SALVe | 1428 | 14 | 17 | 3 | *SCAS | N | Low |
| 13 | HUNT | 1417 | 12-13 | 17-19 | 5 | *HSCL | N | Low |
| 31 | REAL | 1160 | 12-15 | 17-20 | 11 | *MASC | N | Low |
| 5 | NDIT | 781 | 12 | 20 | 8 | CCHS | Y | Low |
| 12 | NANS | 17550 | 12-15 | 13-16 | 1 | *SCAS (Social Anxiety) | Y | Very Low |
| 36 | Raine | 1628 | 8 | 22 | 14 | *DASS-21 | N | Very Low |
| 45 | PSID | 1115 | 12-18 | 18-24 | 6 | *KPDS | N | Very Low |
| 30 | MATCH | 932 | 9-13 | 13-17 | 4 | *PedsQL (Emotional) | N | Very Low |
| 6 | MSLT | 900 | 15 | 24 | 9 | 3-items measuring worry | N | Very Low |
| 51 | MoMo | 686 | 6 | 17 | 11 | *SDQ (Emotional Symptoms) | N | Very Low |
| 94 | n/a (Primary Data) | 558 | 12-17 | 13-18 | 1 | *HADS | N | Very Low |
| 67 | n/a (Primary Data) | 145 | 7-9 | 8-10 | 1 | *SPAIC; *PQSACY; *TRF | Y | Very Low |

**Table 4:**
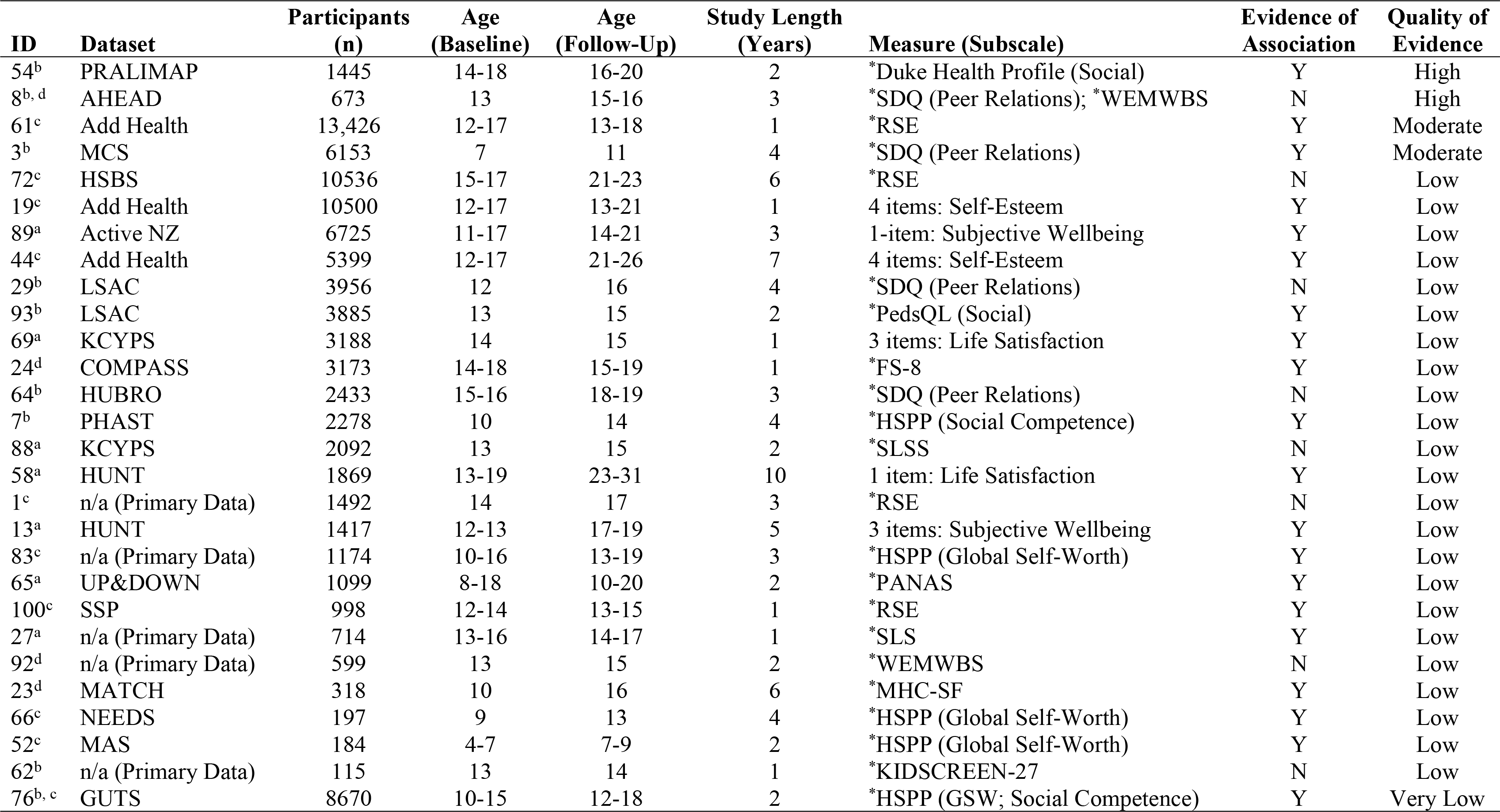
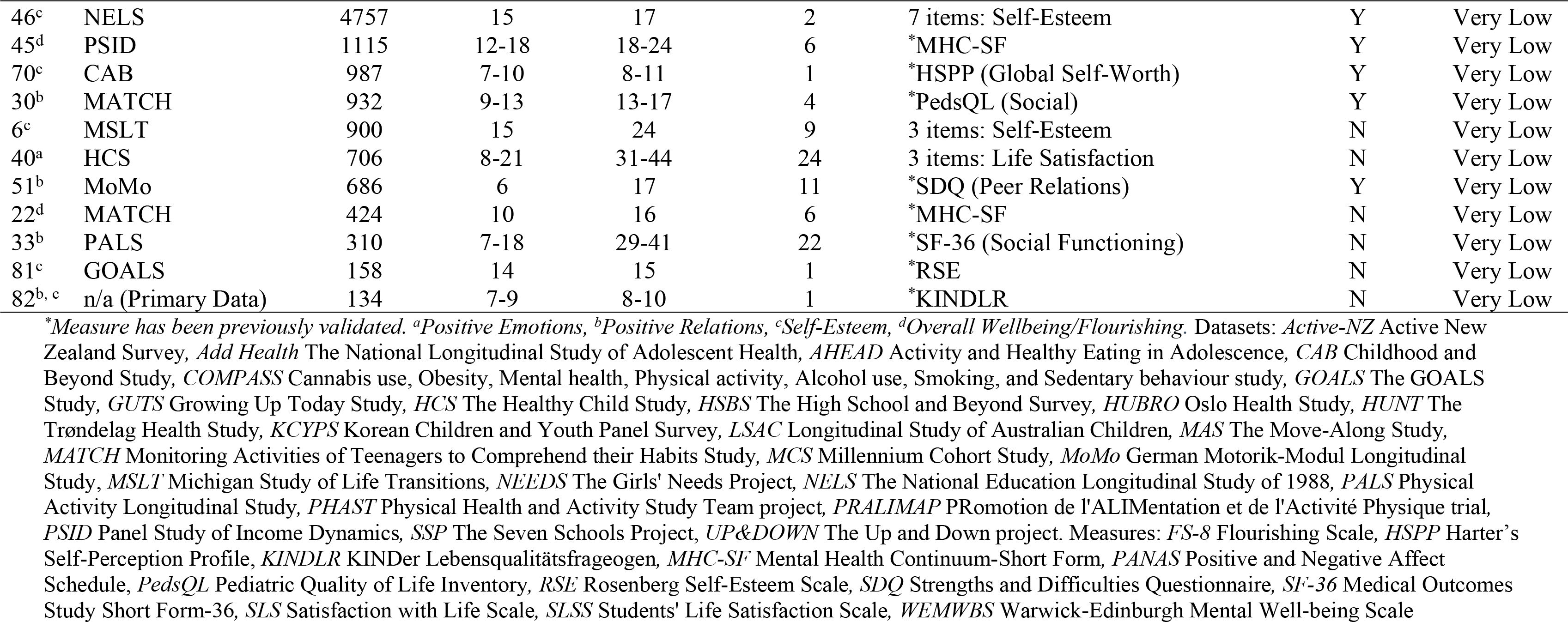
Studies investigating associations between physical activity and indicators of wellbeing

| ID | Dataset | Participants<br>(n) | Age<br>(Baseline) | Age<br>(Follow-Up) | Study Length<br>(Years) | Measure (Subscale) | Evidence of<br>Association | Quality of<br>Evidence |
| --- | --- | --- | --- | --- | --- | --- | --- | --- |
| 54<sup>b</sup> | PRALIMAP | 1445 | 14-18 | 16-20 | 2 | *Duke Health Profile (Social) | Y | High |
| 8<sup>b, d</sup> | AHEAD | 673 | 13 | 15-16 | 3 | *SDQ (Peer Relations); *WEMWBS | N | High |
| 61<sup>c</sup> | Add Health | 13,426 | 12-17 | 13-18 | 1 | *RSE | Y | Moderate |
| 3<sup>b</sup> | MCS | 6153 | 7 | 11 | 4 | *SDQ (Peer Relations) | Y | Moderate |
| 72<sup>c</sup> | HSBS | 10536 | 15-17 | 21-23 | 6 | *RSE | N | Low |
| 19<sup>c</sup> | Add Health | 10500 | 12-17 | 13-21 | 1 | 4 items: Self-Esteem | Y | Low |
| 89<sup>a</sup> | Active NZ | 6725 | 11-17 | 14-21 | 3 | 1-item: Subjective Wellbeing | Y | Low |
| 44<sup>c</sup> | Add Health | 5399 | 12-17 | 21-26 | 7 | 4 items: Self-Esteem | Y | Low |
| 29<sup>b</sup> | LSAC | 3956 | 12 | 16 | 4 | *SDQ (Peer Relations) | N | Low |
| 93<sup>b</sup> | LSAC | 3885 | 13 | 15 | 2 | *PedsQL (Social) | Y | Low |
| 69<sup>a</sup> | KCYPS | 3188 | 14 | 15 | 1 | 3 items: Life Satisfaction | Y | Low |
| 24<sup>d</sup> | COMPASS | 3173 | 14-18 | 15-19 | 1 | *FS-8 | Y | Low |
| 64<sup>b</sup> | HUBRO | 2433 | 15-16 | 18-19 | 3 | *SDQ (Peer Relations) | N | Low |
| 7<sup>b</sup> | PHAST | 2278 | 10 | 14 | 4 | *HSPP (Social Competence) | Y | Low |
| 88<sup>a</sup> | KCYPS | 2092 | 13 | 15 | 2 | *SLSS | N | Low |
| 58<sup>a</sup> | HUNT | 1869 | 13-19 | 23-31 | 10 | 1 item: Life Satisfaction | Y | Low |
| 1<sup>c</sup> | n/a (Primary Data) | 1492 | 14 | 17 | 3 | *RSE | N | Low |
| 13<sup>a</sup> | HUNT | 1417 | 12-13 | 17-19 | 5 | 3 items: Subjective Wellbeing | Y | Low |
| 83<sup>c</sup> | n/a (Primary Data) | 1174 | 10-16 | 13-19 | 3 | *HSPP (Global Self-Worth) | Y | Low |
| 65<sup>a</sup> | UP&DOWN | 1099 | 8-18 | 10-20 | 2 | *PANAS | Y | Low |
| 100<sup>c</sup> | SSP | 998 | 12-14 | 13-15 | 1 | *RSE | Y | Low |
| 27<sup>a</sup> | n/a (Primary Data) | 714 | 13-16 | 14-17 | 1 | *SLS | Y | Low |
| 92<sup>d</sup> | n/a (Primary Data) | 599 | 13 | 15 | 2 | *WEMWBS | N | Low |
| 23<sup>d</sup> | MATCH | 318 | 10 | 16 | 6 | *MHC-SF | Y | Low |
| 66<sup>c</sup> | NEEDS | 197 | 9 | 13 | 4 | *HSPP (Global Self-Worth) | Y | Low |
| 52<sup>c</sup> | MAS | 184 | 4-7 | 7-9 | 2 | *HSPP (Global Self-Worth) | Y | Low |
| 62<sup>b</sup> | n/a (Primary Data) | 115 | 13 | 14 | 1 | *KIDSCREEN-27 | N | Low |
| 76<sup>b, c</sup> | GUTS | 8670 | 10-15 | 12-18 | 2 | *HSPP (GSW; Social Competence) | Y | Very Low |
| 46<sup>c</sup> | NELS | 4757 | 15 | 17 | 2 | 7 items: Self-Esteem | Y | Very Low |
| 45<sup>d</sup> | PSID | 1115 | 12-18 | 18-24 | 6 | *MHC-SF | Y | Very Low |
| 70<sup>c</sup> | CAB | 987 | 7-10 | 8-11 | 1 | *HSPP (Global Self-Worth) | Y | Very Low |
| 30<sup>b</sup> | MATCH | 932 | 9-13 | 13-17 | 4 | *PedsQL (Social) | Y | Very Low |
| 6<sup>c</sup> | MSLT | 900 | 15 | 24 | 9 | 3 items: Self-Esteem | N | Very Low |
| 40<sup>a</sup> | HCS | 706 | 8-21 | 31-44 | 24 | 3 items: Life Satisfaction | N | Very Low |
| 51<sup>b</sup> | MoMo | 686 | 6 | 17 | 11 | *SDQ (Peer Relations) | Y | Very Low |
| 22<sup>d</sup> | MATCH | 424 | 10 | 16 | 6 | *MHC-SF | N | Very Low |
| 33<sup>b</sup> | PALS | 310 | 7-18 | 29-41 | 22 | *SF-36 (Social Functioning) | N | Very Low |
| 81<sup>c</sup> | GOALS | 158 | 14 | 15 | 1 | *RSE | N | Very Low |
| 82<sup>b, c</sup> | n/a (Primary Data) | 134 | 7-9 | 8-10 | 1 | *KINDLR | N | Very Low |

Indicators of wellbeing were mostly measured using subscales of larger questionnaires typically used to quantify a range of outcomes both related and unrelated to mental health. The FS-8 [52], KINDL-R [53], WEMWBS [54] and MHC-SF [10] were the only dedicated wellbeing/flourishing scales that targeted elements of social, emotional and psychological wellbeing simultaneously. These scales were relatively underutilised, appearing in only 7/39 (17.9%) wellbeing studies combined^(8, 22, 23, 24, 45, 82, 92)^. The RSE^(1, 61, 72, 81, 100)^ [55] and/or HSPP Global Self-Worth subscale^(52, 66, 70, 76, 83)^ [56], and the SDQ Peer Relationships subscale^(3, 8, 29, 51, 64)^ [51] were the most common measures of self-esteem (psychological wellbeing) and positive relationships (social wellbeing), respectively. Positive emotions (emotional wellbeing) were only investigated using validated questionnaires in three studies using the SLS^(27)^ [57], SLSS^(88)^ [58] and the PANAS Positive Affect subscale^(65)^ [59]. Additional studies measured emotional wellbeing via (to the best of our knowledge) unvalidated items, so findings should be treated with caution^(13, 40, 58, 69, 89)^.

### Results of synthesis

To address our primary research questions, studies are grouped by mental health outcome into SoF tables structured in order of quality of evidence and sample size (see Tables 2-7). Studies investigating multiple outcomes are reported in all relevant tables. Baseline age represents the age at which PA data was first collected whilst follow-up age represents the age participants last reported on their mental health.

Fig 3 clusters studies by mental health outcome reported, length of follow-up (i.e., from first PA measurement to last mental health measurement) and whether they reported a statistically significant beneficial effect. Across nearly all timeframes and all outcomes, a greater proportion of studies reported a beneficial effect than did not, providing partial evidence that child/adolescent PA may predict anxiety, depression and wellbeing at least 12 months later and as far as 55 years across the lifespan^(90)^.

**Figure 3:**
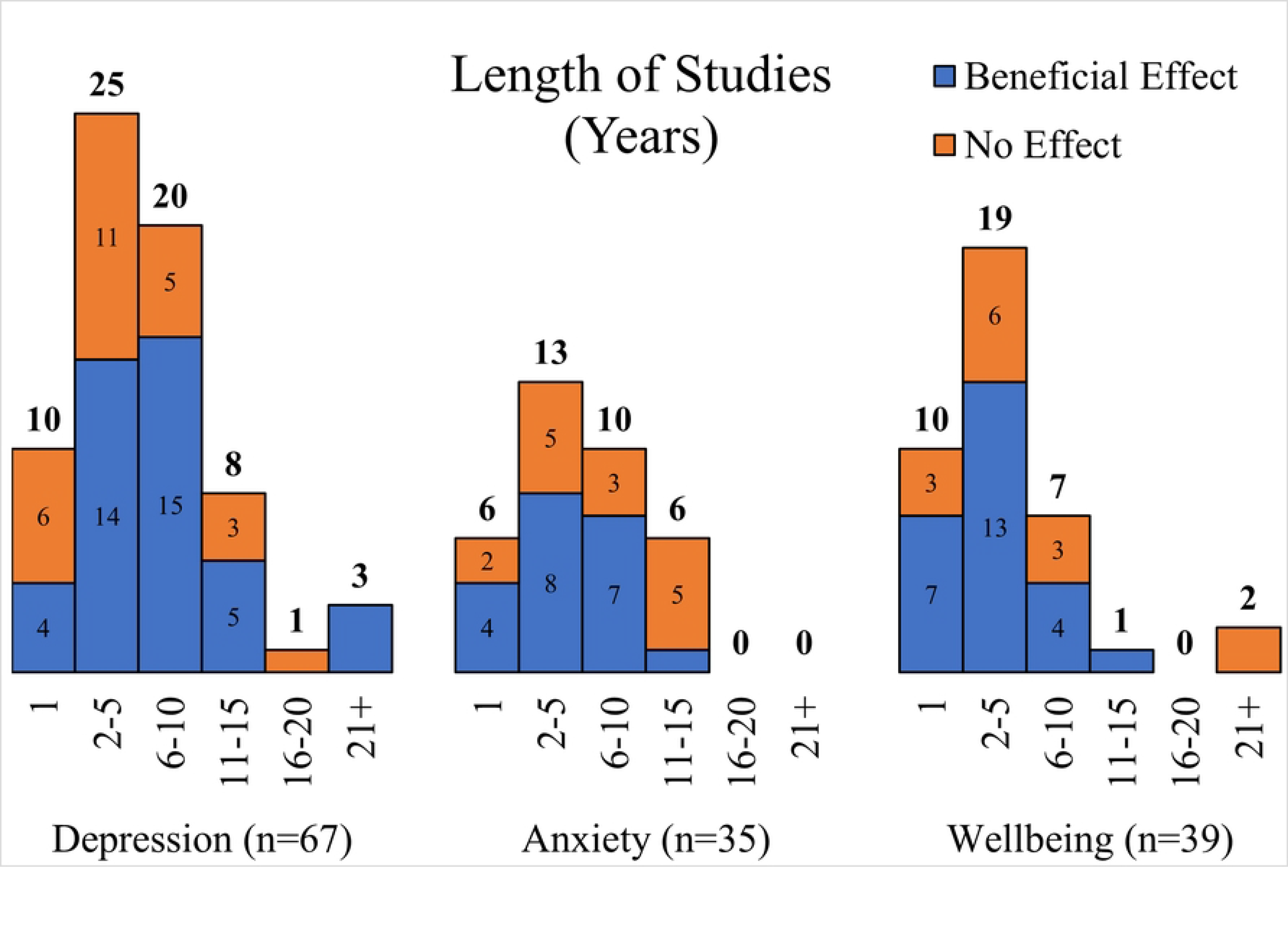
Length of studies in years grouped by outcome

A total of 41/67 depression studies (61.2%) reported significant negative associations (Table 2). Ten of these studies were moderate-or high-quality^(39, 41, 50, 53, 61, 63, 87, 90, 96, 99)^ and observed 1/42 moderate, 17/42 small, and 24/42 non-significant effects (Table 5). In comparison, 19/35 studies (54.3%) reported significant negative associations with anxiety (Table 3). Of these, 6 were moderate-or high-quality^(3, 8, 50, 87, 91, 99)^ and reported 1/34 moderate, 6/34 small and 27/34 non-significant effects (Table 6). Finally, 25/39 studies (64.1%) investigating indicators of wellbeing reported significant positive associations (Table 4). Of these, 4 were moderate-or high-quality^(3, 8, 54, 61)^ and presented evidence of 2/12 moderate, 5/12 small and 5/12 non-significant effects (Table 7). A key is provided next to the study ID in Table 4 to indicate which indicator of wellbeing/flourishing was investigated. Further details on the 88 associations explored in moderate-or high-quality studies (*n* = 14) can be found in Tables 5, 6, 7.

**Table 5:** Effects sizes in moderate- and high-quality depression studies

| ID | Predictor | Outcome | Measure of Effect | Results<br>(unadjusted) | Results<br>(adjusted) | Comments |
| --- | --- | --- | --- | --- | --- | --- |
| 90 | Inactive (ref) vs Active age 7 years | Depressive Symptoms from 23-62 | Hazard Ratio (HR), 95%CI | .85<br>[.68 to 1.08] | .81<br>[.55 to 1.18] | No difference |
|  | Inactive (ref) vs Active age 11 years | Depressive Symptoms from 23-62 | Hazard Ratio (HR), 95%CI | .91<br>[.78 to 1.07] | .96<br>[.75 to 1.22] | No difference |
|  | Inactive (ref) vs Active age 16 years | Depressive Symptoms from 23-62 | Hazard Ratio (HR), 95%CI | .75<br>[.68 to .83] | .73<br>[.62 to .85] | 27% less risk of elevated depressive symptoms |
| 87 | Weekly PA without coach: Never (ref) vs. 1-3 times age 10 (boys) | (Time until) diagnosis of internalizing disorder or age 18 | Hazard Ratio (HR), 95%CI | n/a | .78<br>[.54 to 1.14] | No difference |
|  | Weekly PA without coach: Never (ref) vs. 4+ times age 10 (boys) | (Time until) diagnosis of internalizing disorder or age 18 | Hazard Ratio (HR), 95%CI | n/a | .98<br>[.69 to 1.38] | No difference |
|  | Weekly PA with coach: Never (ref) vs. 1-3 times age 10 (boys) | (Time until) diagnosis of internalizing disorder or age 18 | Hazard Ratio (HR), 95%CI | n/a | .85<br>[.68 to 1.08] | No difference |
|  | Weekly PA with coach: Never (ref) vs. 4+ times age 10 (boys) | (Time until) diagnosis of internalizing disorder or age 18 | Hazard Ratio (HR), 95%CI | n/a | .73<br>[.55 to .98] | 27% less risk of receiving a diagnosis of an internalizing disorder |
|  | Weekly PA without coach: Never (ref) vs. 1-3 times age 10 (girls) | (Time until) diagnosis of internalizing disorder or age 18 | Hazard Ratio (HR), 95%CI | n/a | .80<br>[.61 to 1.06] | No difference |
|  | Weekly PA without coach: Never (ref) vs. 4+ times age 10 (girls) | (Time until) diagnosis of internalizing disorder or age 18 | Hazard Ratio (HR), 95%CI | n/a | .77<br>[.59 to 1.02] | No difference |
|  | Weekly PA with coach: Never (ref) vs. 1-3 times age 10 (girls) | (Time until) diagnosis of internalizing disorder or age 18 | Hazard Ratio (HR), 95%CI | n/a | 1.03<br>[.86 to 1.23] | No difference |
|  | Weekly PA with coach: Never (ref) vs. 4+ times age 10 (girls) | (Time until) diagnosis of internalizing disorder or age 18 | Hazard Ratio (HR), 95%CI | n/a | .92<br>[.71 to 1.18] | No difference |
| 61 | Times engaging in PA in past week during adolescence (12-17) (males) | Depressive Symptoms after one year | Unstandardized Beta (B), (SE), *p* | n/a | -.125 (.027)<br>*p* < .01 | Each additional bout of weekly PA had a small negative effect on depressive symptoms |
|  | Times engaging in PA in past week during adolescence (12-17) (females) | Depressive Symptoms after one year | Unstandardized Beta (B), (SE), *p* | n/a | -.040 (.039)<br>*p* > .10 | No difference |
| 96 | MVPA Guidelines: Didn't Meet (ref) vs. Met in Adolescence (12-17) | Depressive symptoms or disorder age 33-39 | Incidence Rate Ratio (IRR), 95%CI | n/a | .84<br>[.72 to .98] | 16% lower incidence of depression |
|  | MVPA Guidelines: Never Met (ref) vs. Met in Adolescence (12-17) | Depressive symptoms or disorder age 33-39 | Incidence Rate Ratio (IRR), 95%CI | n/a | 1.02<br>[.86 to 1.08] | No difference |
|  | MVPA Guidelines: Never Met (ref) vs. Met in Adulthood (33-39) | Depressive symptoms or disorder age 33-39 | Incidence Rate Ratio (IRR), 95%CI | n/a | .91<br>[.77 to .99] | 9% lower incidence of depression |
|  | MVPA Guidelines: Never Met (ref) vs. Consistently Met (12-17 & 33-39) | Depressive symptoms or disorder age 33-39 | Incidence Rate Ratio (IRR), 95%CI | n/a | .56<br>[.40 to .79] | 44% lower incidence of depression |
| 41 | PA: Total counts age 12 (per 100) | Depressive symptoms age 18 | Incidence Rate Ratio (IRR), 95%CI | .910<br>[.882 to .939] | .941<br>[.910 to .972] | Additional 100 counts of daily activity associated with 5.9% reduction in depression |
|  | PA: Total counts age 14 (per 100) | Depressive symptoms age 18 | Incidence Rate Ratio (IRR), 95%CI | .933<br>[.902 to .965] | .965<br>[.932 to .999] | Additional 100 counts of daily activity associated with 3.5% reduction in depression |
|  | PA: Total counts age 16 (per 100) | Depressive symptoms age 18 | Incidence Rate Ratio (IRR), 95%CI | .939<br>[.896 to .983] | .984<br>[.937 to 1.033] | No difference in fully adjusted model |
|  | Light PA age 12 (per 60mins) | Depressive symptoms age 18 | Incidence Rate Ratio (IRR), 95%CI | .883<br>[.834 to .933] | .904<br>[.850 to .961] | Additional 60mins light activity associated with 9.6% reduction in depression |
|  | Light PA age 14 (per 60mins) | Depressive symptoms age 18 | Incidence Rate Ratio (IRR), 95%CI | .908<br>[.851 to .970] | .922<br>[.857 to .992] | Additional 60mins light activity associated with 7.8% reduction in depression |
|  | Light PA age 16 (per 60mins) | Depressive symptoms age 18 | Incidence Rate Ratio (IRR), 95%CI | .882<br>[.810 to .961] | .889<br>[.809 to .974] | Additional 60mins light activity associated with 11.1% reduction in depression |
|  | MVPA age 12 (per 15mins) | Depressive symptoms age 18 | Incidence Rate Ratio (IRR), 95%CI | .848<br>[.863 to .965] | .910<br>[.857 to .966] | Additional 15mins MVPA associated with 9.0% reduction in depression |
|  | MVPA age 14 (per 15mins) | Depressive symptoms age 18 | Incidence Rate Ratio (IRR), 95%CI | .913<br>[.863 to .965] | .960<br>[.905 to 1.018] | No difference in fully adjusted model |
|  | MVPA age 16 (per 15mins) | Depressive symptoms age 18 | Incidence Rate Ratio (IRR), 95%CI | .938<br>[.883 to .997] | 1.001<br>[.936 to 1.071] | No difference in fully adjusted model |
|  | Total counts: Persistently Low vs. Persistently High from age 12-16 | Depressive symptoms age 18 | Incidence Rate Ratio (IRR), 95%CI | .934<br>[.823 to 1.060] | n/a | No difference |
|  | Light: Persistently Low (ref) vs. Persistently Average from age 12-16 | Depressive symptoms age 18 | Incidence Rate Ratio (IRR), 95%CI | .936<br>[.843 to 1.040] | n/a | No difference |
|  | Light: Persistently Low (ref) vs. Persistently High from age 12-16 | Depressive symptoms age 18 | Incidence Rate Ratio (IRR), 95%CI | .804<br>[.652 to .990] | n/a | Persistently high levels of light activity associated with 19.6% fewer depressive symptoms |
|  | MVPA: Persistently Low (ref) vs. Persistently Above Average from age 12-16 | Depressive symptoms age 18 | Incidence Rate Ratio (IRR), 95%CI | 1.006<br>[.884 to 1.145] | n/a | No difference |
|  | MVPA: Persistently Low (ref) vs. Persistently High from age 12-16 | Depressive symptoms age 18 | Incidence Rate Ratio (IRR), 95%CI | .699<br>[.515 to .950] | n/a | Persistently high levels of MVPA associated with 30.1% fewer depressive symptoms |
| 63 | Years of school sport (team) from age 12-17 | Depressive Symptoms age 20 | Standardized Beta ($\beta$), *p* | n/a | -.09<br>*p* < .05 | Small negative association between years of team sport and depressive symptoms |
| | Years of school sport (individual) from age 12-17 | Depressive Symptoms age 20 | Standardized Beta ($\beta$), $p$ | n/a | .01<br>$p > .05$ | No effect |
| 39 | Years of adolescent school sport (any) from age 12-17 | Depressive Symptoms age 20 | Standardized Beta ($\beta$), $p$ | n/a | -.09<br>$p < .05$ | Years of sport had small negative association with depressive symptoms |
| 53 | Specialist Taught vs. Regular PE age 8: end of 12-month intervention | Change in depressive symptoms age 9 | $p$ | $p > .05$ | n/a | No difference |
| | Specialist Taught vs. Regular PE age 8: after 4 years | Change in depressive symptoms age 12 | $p$ | $p > .05$ | n/a | No difference |
| 99 | All Sport: Low (ref) vs. High trajectory from age 6-10 | Depressive symptoms age 10 | Standardized Beta ($\beta$), 95%CI | n/a | -.10<br>[-.18 to -.02] | Small negative effect of high sport trajectory on depressive symptoms |
| | High Team (ref) vs. High Individual sport participation from age 6-10 | Depressive symptoms age 10 | Standardized Beta ($\beta$), 95%CI | n/a | .05<br>[-.05 to .16] | No difference |
| | Team Sport: Low (ref) vs High trajectory from age 6-10 | Depressive symptoms age 10 | Standardized Beta ($\beta$), 95%CI | n/a | -.12<br>[-.20 to -.03] | Small negative effect of high team sports trajectory on depressive symptoms |
| | Individual Sport: Low (ref) vs. High trajectory from age 6-10 | Depressive symptoms age 10 | Standardized Beta ($\beta$), 95%CI | n/a | -.05<br>[-.13 to .03] | No difference |
| 50 | Team Sport: Non-Participants (ref) vs. Discontinuers from age 12-20 | Depressive Symptoms age 20 | Odds Ratio (OR), 95%CI | n/a | .73<br>[-.66 to 3.53] | No effect |
|  | Team Sport: Non-Participants (ref) vs. Sustainers from age 12-20 | Depressive Symptoms age 20 | Odds Ratio (OR), 95%CI | n/a | .41<br>[-1.30 to 2.13] | No effect |

**Table 6:**
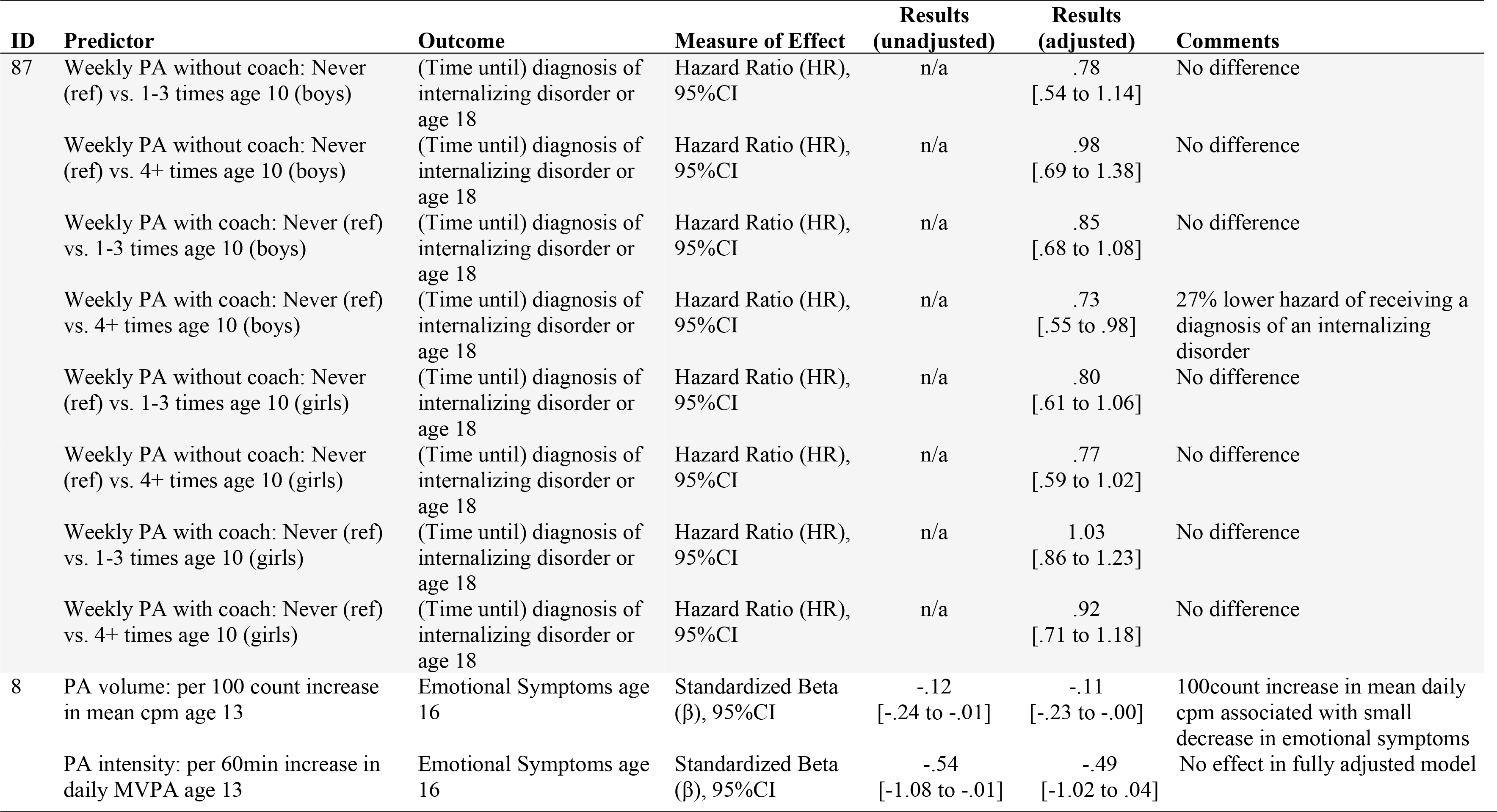
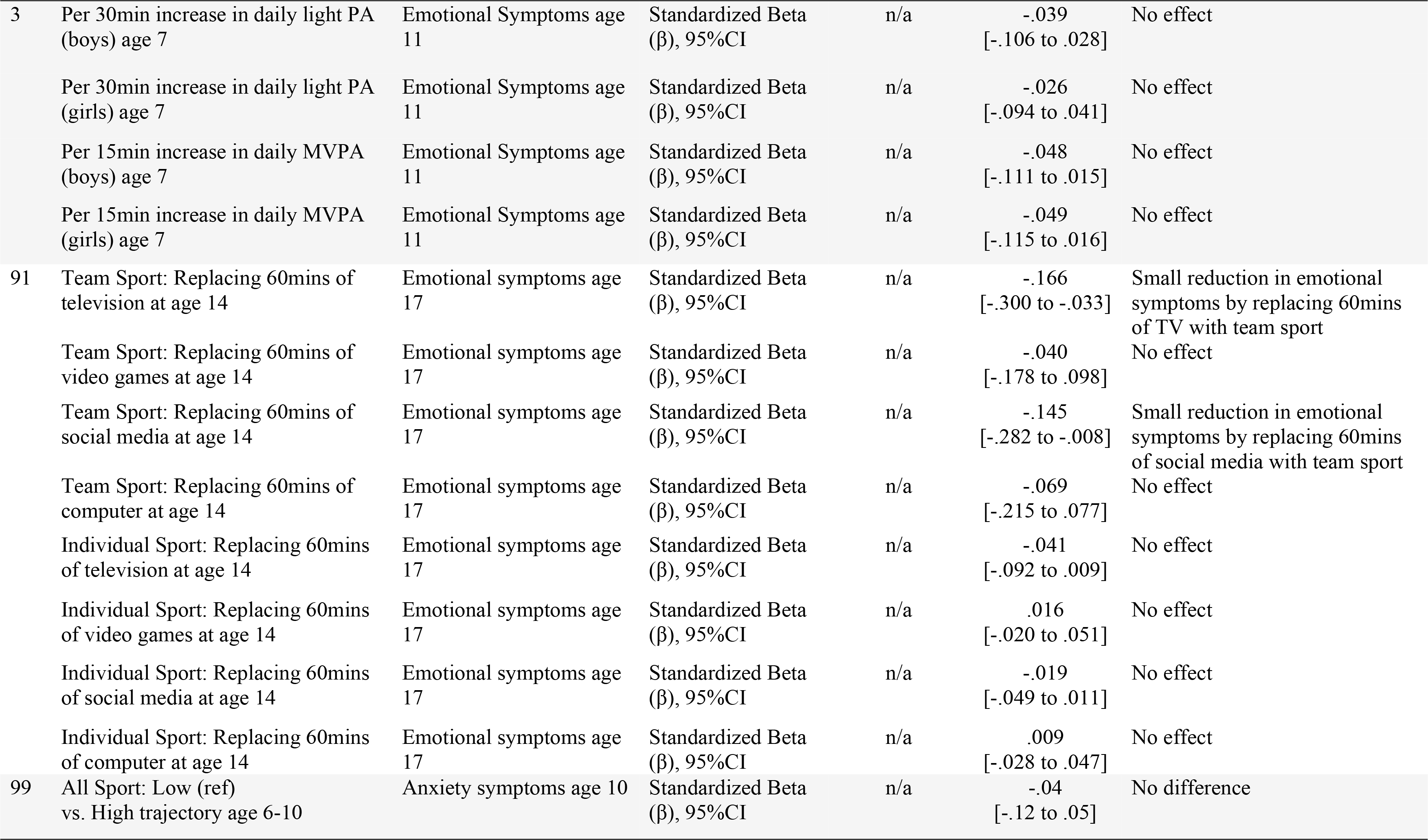
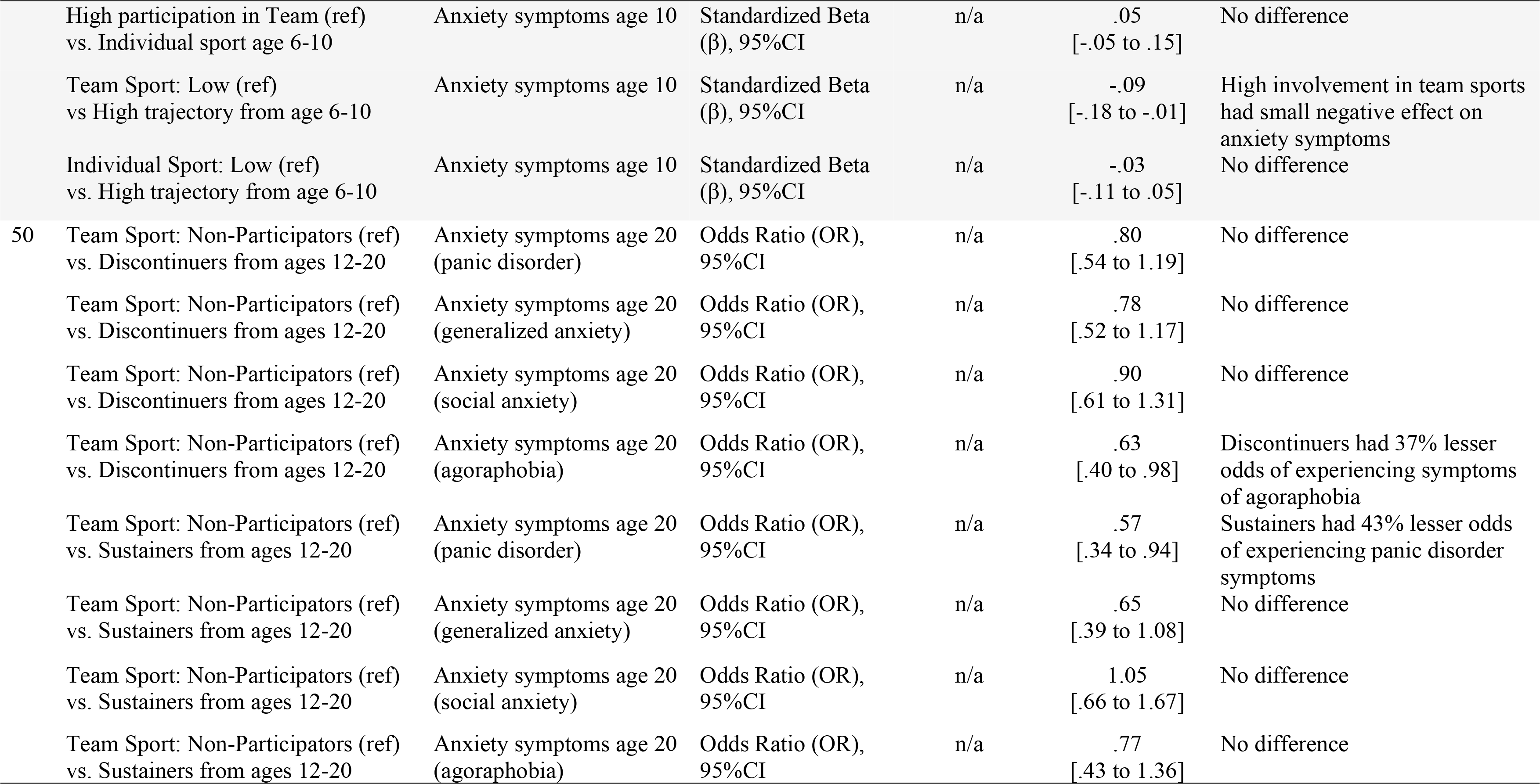
Effects sizes in moderate- and high-quality anxiety studies

**Table 7:** Effects sizes in moderate- and high-quality wellbeing studies

| ID | Predictor | Outcome | Measure of Effect | Results (unadjusted) | Results (adjusted) | Comments |
| --- | --- | --- | --- | --- | --- | --- |
| 54 | Consistently Not Meeting (ref) vs. Meeting WHO PA recommendations one year in 14 to 18-year-olds | Social Health after two years (16-20) | Unstandardized Beta (B), (SE), *p* | n/a | 5.8 (1.3)<br>*p* < .0001 | Meeting guidelines at one timepoint associated with "clinically meaningful" improvement in social health |
|  | Consistently Not Meeting (ref) vs. Meeting WHO PA recommendations both years in 14 to 18-year-olds | Social Health after two years (16-20) | Unstandardized Beta (B), (SE), *p* | n/a | 9.3 (1.2)<br>*p* < .0001 | Consistently meeting guidelines associated with a moderate, "clinically meaningful" improvement in social health |
| 8 | PA volume: per 100 count increase in mean counts per minute age 13 | Wellbeing age 16 | Standardized Beta (β), 95%CI | -.06<br>[-.47 to .35] | -.02<br>[-.44 to .40] | No effect in fully adjusted model |
|  | PA intensity: per 60min increase in daily MVPA age 13 | Wellbeing age 16 | Standardized Beta (β), 95%CI | -.54<br>[-2.49 to 1.41] | .03<br>[-1.98 to 2.05] | No effect in fully adjusted model |
|  | PA volume: per 100 count increase in mean counts per minute age 13 | Peer Relations age 16 | Standardized Beta (β), 95%CI | -.02<br>[-.09 to .06] | -.02<br>[-.09 to .05] | No effect |
|  | PA intensity: per 60min increase in daily MVPA age 13 | Peer Relations age 16 | Standardized Beta (β), 95%CI | -.15<br>[-.49 to .20] | -.16<br>[-.51 to .19] | No effect |
| 61 | Times engaging in PA in past week in 12 to 17-year-olds (males) | Self-Esteem after one year (13-18) | Unstandardized Beta (B), SE, *p* | n/a | .063 (.014)<br>*p* < .01 | Each additional bout of weekly PA had a small positive effect for self-esteem one year later |
|  | Times engaging in PA in past week in 12 to 17-year-olds (females) | Self-Esteem after one year (13-18) | Unstandardized Beta (B), SE, *p* | n/a | .062 (.015)<br>*p* < .01 | Each additional bout of weekly PA had a small positive effect for self-esteem one year later |
| 3 | Per 30min increase in daily light PA (boys) age 7 | Peer Relations age 11 | Standardized Beta (β), 95%CI | n/a | -.086<br>[-.150 to -.021] | Small reduction in peer relationship problems |
|  | Per 30min increase in daily light PA (girls) age 7 | Peer Relations age 11 | Standardized Beta (β), 95%CI | n/a | -.071<br>[-.130 to -.013] | Small reduction in peer relationship problems |
|  | Per 15min increase in daily MVPA (boys) age 7 | Peer Relations age 11 | Standardized Beta (β), 95%CI | n/a | -.077<br>[-.133 to -.022] | Small reduction in peer relationship problems |
|  | Per 15min increase in daily MVPA (girls) age 7 | Peer Relations age 11 | Standardized Beta (β), 95%CI | n/a | -.007<br>[-.057 to .071] | No effect |

### Individual vs. group-based activities

With just one exception^(53)^, all studies researching the effects of group-based or individual activities investigated sport participation. Team sports were found to be beneficial in 7/13 depression studies (53.8%), 8/9 anxiety studies (88.9%) and 1/2 wellbeing studies (50%). Individual sports were beneficial in 1/8 depression studies (12.5%), 4/6 anxiety studies (66.7%) and 0/1 wellbeing studies (0%). Effect sizes were consistently reported as small on all but two occasions^(25, 50)^ wherein, compared to no participation, involvement in team sport throughout adolescence^(25)^ with sustained participation in early adulthood^(50)^ was associated with 24% lower odds of depressive disorder diagnosis^(25)^, 30% lower odds of anxiety disorder diagnosis^(25)^, and 43% lower odds of panic disorder symptoms^(50)^ in early adulthood. Moderate/high-quality studies reported that team but not individual sport participation in childhood/adolescence had small protective effects against prospective depressive and anxiety symptoms up to 3 years later^(50, 63, 91, 99)^. For further information on study lengths see Fig 4. For further information on all studies targeting either individual or group-based activities see Supporting Table S3.

**Figure 4:**
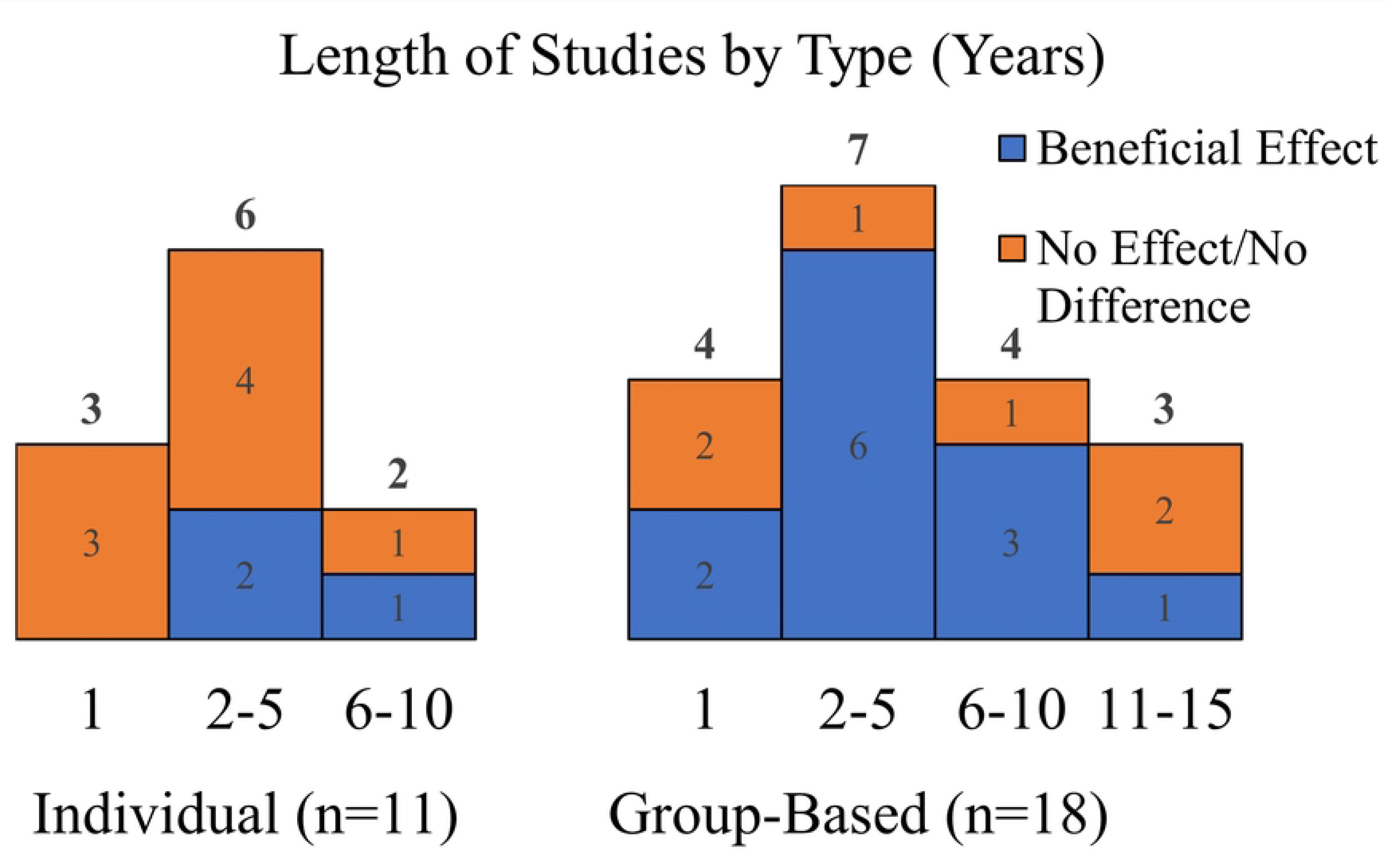
Length of studies in years grouped by type of activity

### Domains of activity

No studies focused on the effects of travel or household activities. In total, leisure-time activities were found to be beneficial in 7/12 depression studies (58.3%), 3/6 anxiety studies (50%) and 5/7 wellbeing studies (71.4%). School-based activities were beneficial in 5/8 depression studies (62.5%), 2/5 anxiety studies (40%) and 2/5 wellbeing studies (40%). Three studies reported moderate effects for depression, anxiety and wellbeing when leisure activity participation was high in adolescence and maintained into adulthood^(47, 50, 58)^. Specifically, 65% reduced relative risk of depression^(47)^, 47% reduced odds of panic disorder symptoms^(50)^, and twice the likelihood high life satisfaction^(58)^ in early adulthood. Moderate and high-quality studies presented evidence that small beneficial effects of school sports (but not PE)^(53)^ and leisure activities for anxiety and depressive symptoms were still observable after 3 years^(50)^ and up to 55 years^(90)^ in the future. For further information on study lengths see Fig 5. For further information on the effects of PA performed in leisure-or school-time see Supporting Table S4.

**Figure 5:**
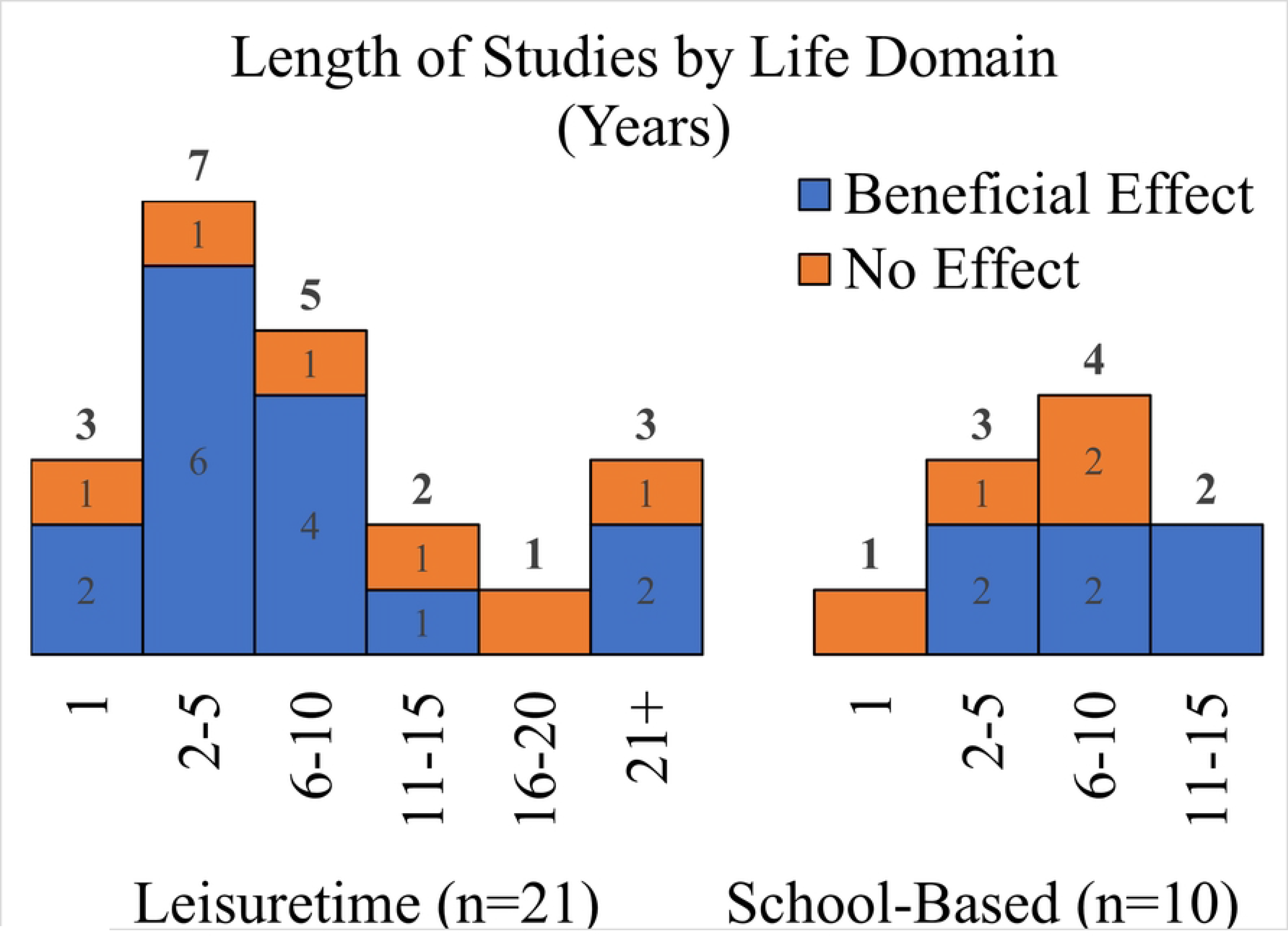
Length of studies in years grouped by domain of activity

### WHO MVPA guideline adherence

In total, 12 studies used data referring to the mean daily amount of MVPA performed across the past 7 days (hence facilitating categorisation) or directly compared the effects of guideline adherence versus non-adherence. Positive effects of guideline adherence were reported in 2/4 depression studies (50%), 1/3 anxiety studies (33.3%) and 4/5 wellbeing studies (80%). Findings were mixed across studies comparing adherence and non-adherence with 1/3 (33.3%) finding additional benefits of achieving meeting guidelines for future depressive symptoms, 0/2 (0%) for anxiety symptoms and 2/2 (100%) for wellbeing-related outcomes. All significant effect sizes reported were small. One study was high quality^(54)^, presenting evidence of a dose response relationship between years of WHO MVPA guideline adherence in adolescence and social quality of life measured one year later. For further information on study lengths see Fig 6. For further evidence of the prospective effects of WHO MVPA guideline (non)adherence, see Supporting Table S5.

**Figure 6:**
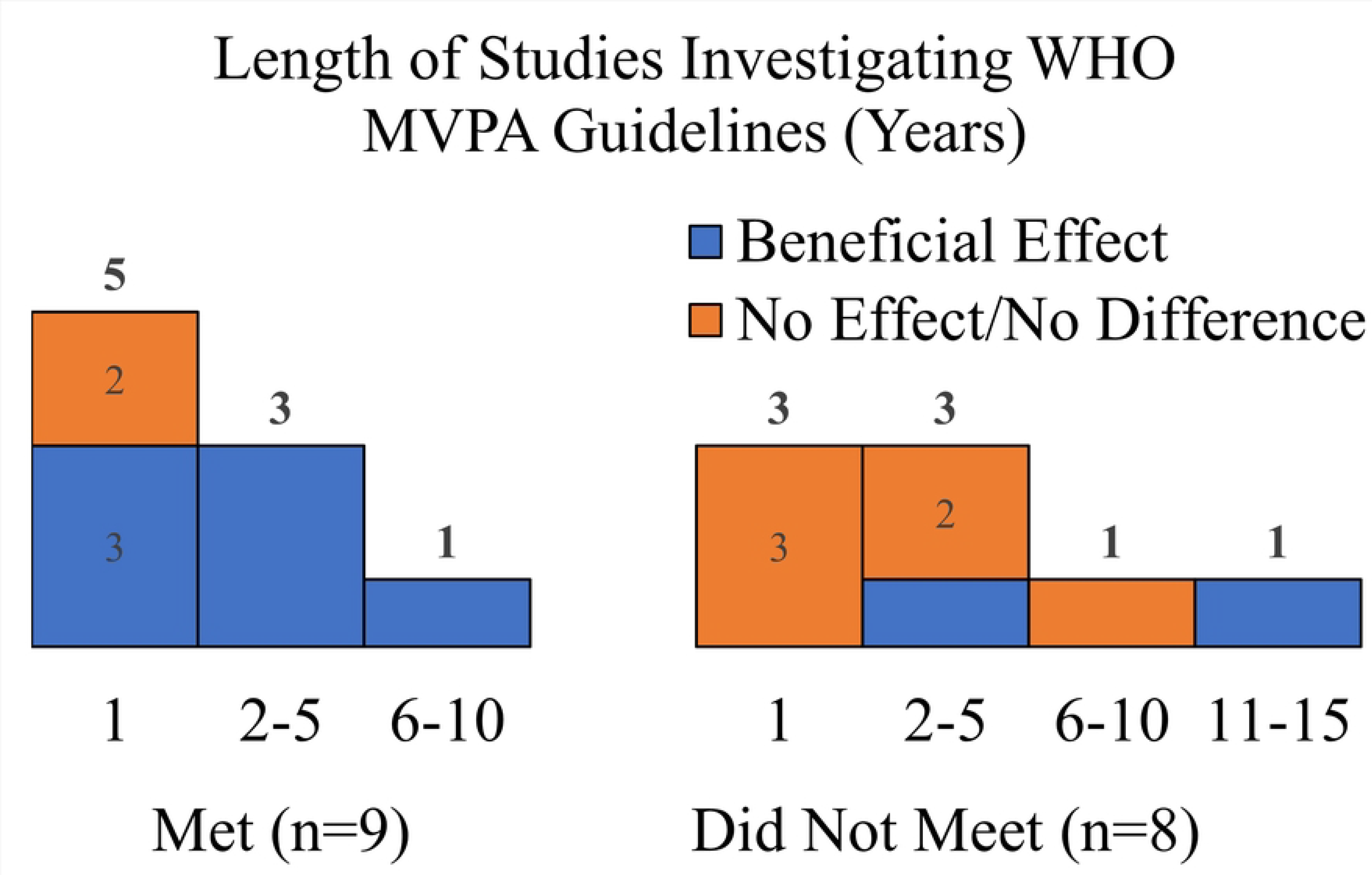
Length of studies in years grouped by WHO MVPA guideline adherence

## Discussion

The present study was the first comprehensive systematic review to assess the association between PA performed in childhood/adolescence for future anxiety, depression and wellbeing using a longitudinal one-year time interval. Evidence of associations with anxiety, depression and wellbeing was deemed to be low quality due to heavy reliance on observational study designs. Moderate and high-quality studies reported small or non-significant effects of child/adolescent PA persisted for at least 12 months suggesting there may be some minor long term mental health benefits that emerge from participation in certain activities in youth. Directing PA promotion towards youth as a method of reducing long-term risk of mental illness or suboptimal wellbeing likely requires careful consideration of important contextual factors in order to be successful. For example, the longest running study in the review was high quality and investigated associations in 18,555 participants over 55 years^(90)^. PA performed age 16 but not age 7 or 11 had protective effects against depressive symptoms that grew in magnitude as participants progressed through mid-life and into old age. This study supports existing literature suggesting that the relationship between PA and prospective mental health outcomes may not be uniformly linear [23]. Inherent complexity in the relationship may be somewhat explained through consideration of type, domain and volume of the activity in question.

### Individual vs. group-based activities

Psychosocial influences can play a key role in the relationship between PA and mental health. In PA research and in particular sports studies, psychosocial factors are commonly investigated by drawing comparisons between individual and group-based activities. Currently, the general hypothesis is that group-based or team-sport environments are more conducive to the development of interpersonal connections and social support networks than individual ones [36],[37]. The current review supports this stance with evidence that team sports are more consistently beneficial for long-term mental health outcomes than individual sports [60]. It is possible that group-based environments offer more opportunities for positive interpersonal interactions than individual environments, thus fulfilling an individual’s need for relatedness and by extension, social wellbeing [61]. In addition, the readily available and naturally forming social support networks, social identification, and sense of belonging that can emerge through team membership may play a role in the reduction of depressive and anxiety symptoms [62].

Included studies investigated a number of psychosociological factors that influence the association between sport participation and mental health. Two studies controlled for mediating effects of sport self-concept and perceived sporting competence^(70, 83)^ reporting that links with future self-esteem were fully dependent on positive sport-related self-beliefs which aligns with existing literature [62]. These studies, among others, present important points for PA promotion. Long-term mental health benefits of sport may not be felt equally for all individuals [60]. For many, PA, whether accumulated through physical education, school sport, or otherwise, can be perceived as a chore and even unpleasant [63],[64] in which case, positive mental health outcomes are likely to be limited. Activities that can take place outside of competitive environments or those that do not lend themselves as easily to comparison with others may be more suitable for individuals with negative self-perceptions, at least in the first instance.

There are also some risks with sport involvement that were not addressed in the review such as the young person’s goal-orientation or athletic identity. Goal Achievement Theory posits that ego-orientated goals can negatively impact wellbeing when playing poorly [65]. Likewise, strong athletic identities have been linked with anxiety during non-normative transitions [66]. It is important to be mindful of these potentially harmful aspects of participation throughout PA promotion and intervention design.

### Domains of activity

Studies in the current review assessed only the effects of leisure or school-based activities leaving the prospective effects of active transport and household activities in young people to be determined. Whether there are differential effects associated with leisure or school-based activities in childhood/adolescence for prospective mental health outcomes remains unclear. One high-quality study^(90)^ found no association between leisure activities performed age 7 or 11 while those performed at age 16 were associated with a 27% reduced risk of depressive symptoms between the ages of 23-62. This could infer the existence of a critical period during which being active yields greater long term mental health effects. Alternatively, such an outcome could be a product of the significant drop off in activity levels that typically coincides the transition from childhood to adolescence [67]. If activity levels reported age 7 or 11 did not track well over time they are less likely to predict future mental health [27].

Two moderate quality studies tested associations with school-based activities^(39, 53)^. One reported that participating in school sports in adolescence had a small negative association with depressive symptoms in early adulthood^(39)^ whilst the other reported that specialist taught PE classes at age 8 had no effect on depressive symptoms after 4 years and was even associated with less decline in symptoms compared to controls^(53)^. Again, inconsistent effects reported by these studies could be linked to poor tracking of PA levels across the transition from childhood to adolescence. They may also illuminate the importance of intrinsic motivation to participate which was more likely a contributing factor for those playing extracurricular sports than in compulsory PE classes [61].

Many explanatory mechanisms for domain-specific effects are rooted in self-determination theory [61]. Research in adults would suggest that household activities are seldom challenging enough to experience a sense of task-mastery, nor are they often fuelled by intrinsic motivation, and hence may not benefit mental health [38]. Indeed, in certain circumstances, these activities might even cause stress. In contrast, it is plausible that consistently connecting with the natural environment by walking to school with friends coupled with intrinsic motivation to be active could simultaneously satisfy one’s need to connect with nature and each other [68],[69]. Ease of access to green space is cited alongside increased surveillance of life domains in the WHOs Global Action Plan for PA [20]. For these reasons, more research on domain-specific PA, particularly active transport is required.

In adults, leisure and transport activities are more likely to be autonomy-supportive and provide opportunities to experience a sense of belonging than occupational or household activities [70]. Whilst adult-based literature can be used to inform hypotheses in children/adolescents, researchers should be careful not to generalise findings across these groups. Domain-specific effects on wellbeing can vary across age ranges with evidence that active transport is beneficial for adults but not adolescents [38]. Benefits may materialise for adults through autonomous endorsement of transport PA for enjoyment or health reasons compared to young people who may have to use active means of transport due to lack of an alternative [38]. Improving understanding of domains could help individuals working in public health develop more strategic interventions best suited to promote PA and mental health in different age groups and communities.

### WHO MVPA guideline adherence

A moderate-quality study using the National Longitudinal Study of Adolescent Health (Add Health) data found that adherence vs. non-adherence to WHO MVPA guidelines in adolescence was associated with a 16% reduction in the incidence of depressive symptoms when aged 33 to 39^(96)^. Trajectory analysis from the same study found evidence of an added benefit for those who continuously met guidelines (i.e., throughout adolescence and adulthood) compared those who started to meet guidelines later in life. Whilst findings do not dispute the commonly held belief that it is never too late to start being active, they do infer that the younger we start, the better.

Guidelines dictate that across the week, children and adolescents should engage in an average of 60 minutes MVPA per day to experience health benefits. It is important to note that current MVPA guidelines are not specific to mental health and also aim to support physical health through cardiorespiratory fitness [39]. As such, focusing on the effects of 60 minutes of MVPA per day could mean that benefits associated with lesser amounts of PA are overlooked. Whilst in the current review beneficial effects emerged from guideline adherence, there were several incidences where benefits were achieved through lesser amounts^(30, 51)^ or where there were no differences between those who met and did not meet guidelines^(55, 85)^. The current review presents partial evidence that WHO MVPA guideline adherence may be beneficial for long-term mental health. However, given the inconsistency of findings, perhaps a more realistic or achievable goal would be for young people to simply ‘get moving’. This approach aligns with a growing body of evidence that any activity is better than no activity [71],[72].

Research should continue to explore the benefits of WHO MVPA guideline adherence but broaden its focus to also consider the long-term effects of lesser volumes. Some low-quality studies reported that light-intensity PA performed in childhood/adolescence was associated with better social wellbeing^(3)^ and fewer prospective depressive/anxiety symptoms^(8, 41, 42)^. Increasing light-intensity activity or introducing small bouts of MVPA through programmes such as ‘Snacktivity’ [73] aligns with the small-change approach to habit-formation [74] and may present a more achievable goal for particularly inactive young people. Additionally, PA guidelines for children have only recently begun to incorporate muscle-strengthening activities (e.g., bodyweight exercises, resistance training) into recommendations. In our review, few studies measured such behaviours in young people^(16, 26)^. Given emerging research in adults showing the benefits of muscle strengthening activities for mental health beyond those associated with aerobic PA [75],[76], closer inspection of their association in children/adolescents’ mental health is warranted.

### Heterogeneity & other observations

Heterogeneity in the items used to measure mental health limited the comparability of study findings. For example, aside from two studies using the SMFQ to measure depression^(29, 93)^, every study investigating the effects of individual or group-based activities used a different measure of mental health. In total, 23 validated measures of depression, 19 validated measures of anxiety, and 15 validated measures of wellbeing were used across the 98 studies alongside a plethora of non-validated items. Some scales have been validated for use in the general population however, to the best of our knowledge are yet to undergo formal validation in child/adolescent samples. In attempting to advance the field, psychometric validation studies should assess a range of metrics specifically designed for use in young people for concurrent/divergent validity, test-retest reliability, factor structure, and item comprehension. Such examples exist for health-related quality of life measures in children (e.g., KIDSCREEN-27)[77] but few have undergone the same rigorous assessments for mental illness.

Conversely, depression and anxiety are both unitary constructs with widely accepted definitions, so there is a greater likelihood that different scales converge on the same latent construct than for an ill-defined concept such as wellbeing. Two high quality studies with similar aged participants reported inconsistent effects between comparable levels of daily MVPA and social wellbeing^(8, 54)^. These studies are said to be measuring two broadly similar constructs in social quality of life and peer relationships, but the lack of conceptual clarity makes it difficult to determine if they were comparing like for like. Concurrent validity studies have attempted to address this issue in the past [78] but up-to-date comparisons of wellbeing measures are scant. Rigorous validation studies for wellbeing measures in younger populations are necessary to improve study comparability going forward.

Historical shifts in research focus were observed. Studies using datasets commencing prior to the year 2000 displayed a clear tendency to investigate depression over anxiety and indicators of wellbeing. It could be argued that to reduce the subsequent disbalance in the literature, wellbeing and/or anxiety should be the focus of research going forward. However, technological advancements and ‘The Physical Activity Transition’ [79] toward more sedentary-type jobs/pastimes have given rise to a generational shift in PA patterns. This shift means the relationship between PA and mental health has evolved over time [63] making older data less representative of modern-day associations. PA participation now offers a way to reduce total screentime; an increasingly prevalent lifestyle factor independently linked to depression, anxiety, and self-esteem in young people that did not exist prior to the invention of smart phones [80]. Research must continue to investigate the extent to which PA can influence both mental illness and wellbeing to maintain up-to-date understanding of these perpetually fluid relationships.

### Limitations of included studies

Due to heavy reliance on longitudinal cohort studies, most evidence was low quality. The resources required to monitor the effects of PA at one timepoint for mental health outcomes at least one year later are considerable, meaning the reliance on longitudinal cohort studies and other observational designs is likely to continue in the future. Whilst there is a need for more randomized trials, a more pertinent recommendation would be for those overseeing the management of longitudinal cohort studies to come to a consensus on a theoretically informed understanding of mental health and the measurement tools they use to improve physical activity and mental health epistemology in young people. Such a move would reduce some limitations associated with secondary analysis and enhance study comparability whilst accommodating newly emerging high-level forms of statistical analyses such as retrospective data harmonization [81].

### Limitations of the review design

Several studies used the same data-source to answer similar research questions, posing the risk that associations (or lack thereof) from a single data-source could skew findings [82]. A systematic process was followed [49] to compare studies using the same data-source on a number of study modifiers (see supporting files). Except for two studies^(74, 86)^, those included in data synthesis were all deemed to provide a unique insight into the association between child/adolescent PA and outcomes related to anxiety, depression or wellbeing at least 12 months later (Supporting Table S1). Further, best efforts have been made to ensure the source of observed associations is clear with SoF tables noting the dataset used in each article.

Aligning with Keyes’ two-continua model meant that whilst the review was structured within a sound theoretical framework, a small number of studies had to be excluded as they analysed psychometric scales that combined aspects of mental illness and wellbeing into an overall mental health score. It is the authors’ belief that the increased heterogeneity that would have arisen through inclusion of these additional measures would have obfuscated trends further and that the benefits of using Keyes’ model as a conceptual framework outweighed its drawbacks, strengthening the quality of the review as a whole.

## Conclusions

PA promotion in young people may represent a viable means of achieving a small reduction in the overall burden of CMDs and improving indicators of wellbeing in some individuals however, research is disjointed. The current review presents low quality partial evidence for PA participation in childhood/adolescence having small benefits for mental health outcomes related to anxiety, depression and wellbeing at least 12 months later. Team sport participation is among the most consistently predictive type of PA. Long-term effects of domain-specific activities require further research with particular focus needed on the effects of active transport and household activities. Adhering to WHO MVPA guidelines in youth may be associated with small improvements in long-term mental health however, the benefits of lesser volumes and intensities of activity warrant further discussion. A vast range mental health measures used across studies led to heterogeneous operational definitions and conceptual ambiguity. Researchers must strive to achieve consensus on their theoretical approach and instruments used to measure depression, anxiety, and wellbeing in young people to overcome epistemological concerns and improve study comparability going forward.

## Data Availability

The complete data extraction file has been made available on Figshare: https://doi.org/10.6084/m9.figshare.23580423.v1

## Acknowledgements

We would like to thank all authors who provided additional information used for synthesis.

## Supporting information

**S1 Table: References of all studies included in the review**

**S2 File: Study modifiers**

**S3 Table: Duplicated datasets**

**S4 Table: Study characteristics**

**S5 Table: Studies of individual and group-based physical activity**

**S6 Table: Studies of leisure-time and school-based physical activity**

**S7 Table: Studies investigating effects of WHO MVPA guideline adherence**

**S8 File: Abbreviations for datasets and measures used**

**S9 File: Database search terms S10 File: Review protocol**

